# Body Clock: Matching Personalized Multimorbidity and Fast Aging Using Information Entropy

**DOI:** 10.1101/2021.03.29.21254372

**Authors:** Shabnam Salimi, Aki Vehtari, Marcel Salive, Luigi Ferrucci

## Abstract

With aging, most older adults are at risk of having more than two diseases, conventionally defined as multimorbidity. We determined body organ disease number (BODN) as a new multimorbidity index. We measured the degree to which each disease level, from mild to severe, predicts longitudinal BODN uncoupled from chronological age. We determined Body Clock using global disease levels burden from all systems predicting longitudinal BODN for each individual, which is a proxy of the personalized rate of biological aging. Change in Body Clock predicts late-life age-related outcomes and can be used for geriatric clinics and clinical trials for precision medicine.

---

The **c**urrent medical practice focuses primarily on one disease or on diseases that are coexistent in the same organ system. However, epidemiological data show that multimorbidity, conventionally defined as having two or more diseases, is the most frequent health condition in persons 65 and older ^1–4^. Multimorbidity is associated with a high risk of polypharmacy, iatrogenesis, and geriatric syndromes that imposes burdens on individuals and society ^5^. The rate of aging itself is posited as the underlying cause of pathologies across body systems, with damage accumulation exceeding homeostatic mechanisms and physiological resilience, emerging clinically as multimorbidity ^6, 7^.

The central hypothesis that multiple chronic diseases represent a clinical manifestation of the rate of aging has led to the development of tools for measuring multimorbidity.

Some of these tools consider the number of chronic diseases ^8^, whereas others weigh the contribution of specific diseases based on their estimated impact on functional status ^9^ or mortality ^10, 11^. The first type of tool fails to consider that chronic diseases are highly heterogeneous. The second type can underestimate the health burdens in patients of one potentially fatal disease or fail to recognize individuals can accumulate multiple diseases and still live long lives. Consistent dynamic compensatory mechanisms might counteract specific organ systems’ dysfunctions, while global physical and cognitive functions remain preserved ^9^.

Alternative approaches have estimated the rate of aging using indices that aggregate multiple physiological and/or blood biomarkers ^12–14^. In most cases, chronological age (c-age) to predict mortality or self-reported health ^12, 14, 15^ or late-life outcomes ^16, 17^ were included as part of the indices. Including c-age or late-life outcomes might skew the metric toward older c-ages and fail to capture specific early health deterioration. Moreover, indices based on volatile biomarkers might be prone to fluctuations due to transient stress levels and entropic forces, not distinguished between damage and adaptation, masking underlying pathophysiological changes related to multimorbidity.

An ideal tool should capture sub-clinical to clinical states as an overarching health assessment and address the complexity of multimorbidity uncoupled from c-age and independent of late-life outcomes ^18, 19^. With these characteristics in mind, we developed a global outcome measure of multimorbidity called *Body Organ Disease Number* (BODN) as an ordinal number ranging 0-13. BODN is the number of organ systems with at least one disease. We assessed the beta coefficient estimates of all diseases at two lagged times that incorporate into BODN over time uncoupled from c-age. Given the premise of interindividual variability in rates of chronic disease manifestation and using all disease levels in a model, we predicted BODN over time using lagged disease-levels burdens at individual-levels as the personalized rate of biological aging termed *Body Clock*.

We used Bayesian inference to obtain maximum information on the coefficient estimates and incorporated health deterioration from subclinical to clinically diagnosed diseases. We show that Body Clock can predict late-life outcomes ranging from physical functional declines to death.

## Results

To calculate observed BODN, the presence of a specific disease for each organ system was established based on predefined impairment criteria as either deviation from values at healthy young or commonly accepted diagnoses in clinical practice. A specific organ system with at least one positive disease contributes to the BODN as a global multimorbidity measure. In addition to 11 organ systems, we also counted cerebrovascular accident (CVA), sharing pathophysiology across several organ systems, and cancer, with its distinct pathophysiology, giving BODN a range from 0 to 13 as an ordinal outcome (Fig. 1a). Over the study period, median BODN increased with age so that for those at age 27-44 years, BODN was 2 (range:0-8), at 45-54 years was 3 (range 0-11), at 55-64 was 4 (range0-11), at 65-74 was 5 (range: 2-11), at 74-85 was 6 (range: 2-11), at older than 85 was 7 (range: 3-12). As lagged predictors, the organ-specific diseases were assigned an ordinal number for their deviation from younger age, severity or stage of disease (Table 1).

**Figure 1.**
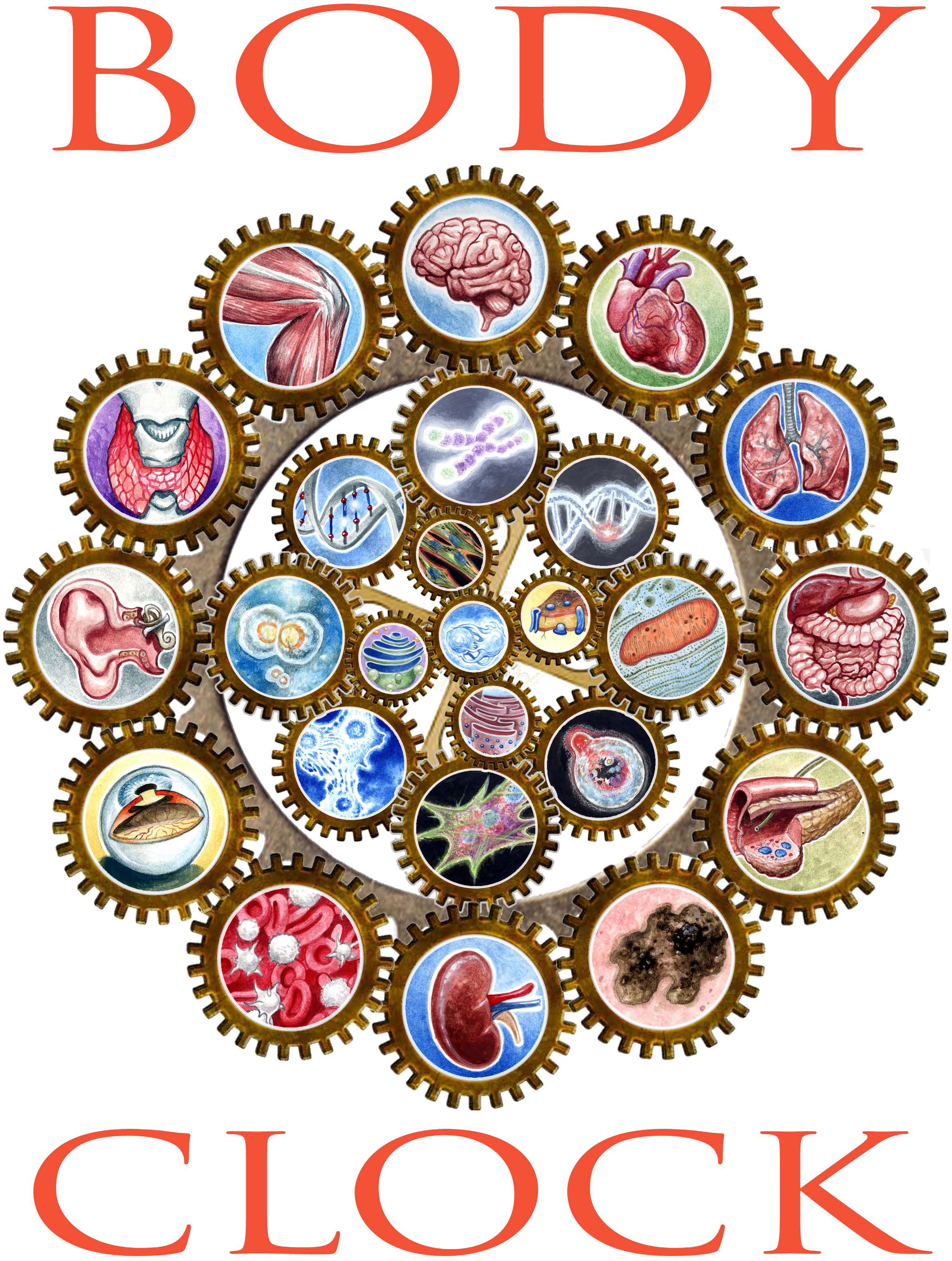
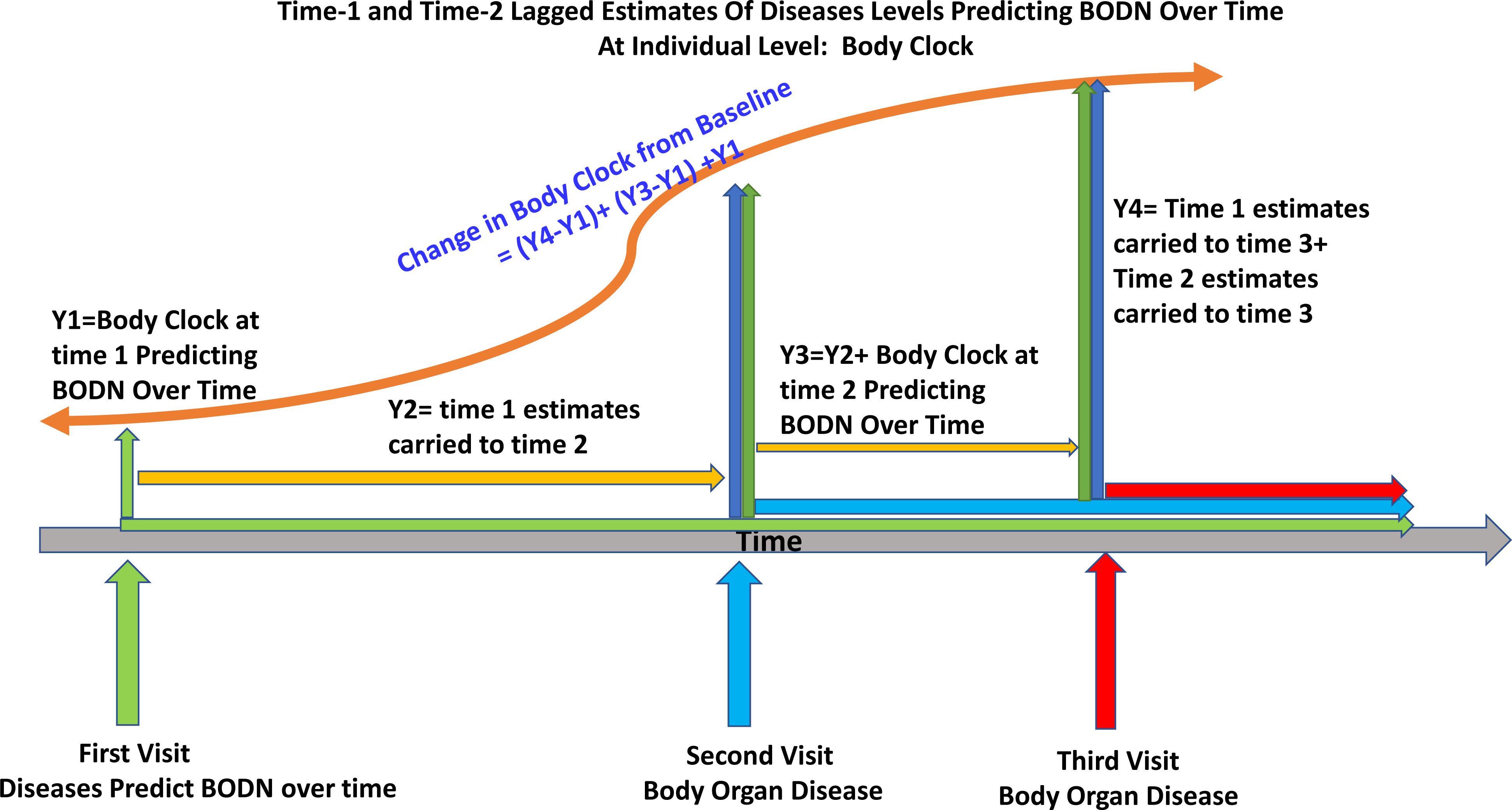
**(a)** Symbolic representation of the integrated system-based calculated Body Clock Body organ disease number (BODN) considers 11 organs and two diseases, cerebrovascular accidents and cancer. The predicted BODN at an individual level is quantified, termed Body Clock. Clockwise, the Body Clock includes the central nervous system and cerebrovascular accidents, cardiovascular system, respiratory system, gastrointestinal and liver system, metabolic system, cancer, renal system, hematopoietic system, the sensory systems (ophthalmic and hearing), dysthyroidism, and musculoskeletal system. **(b)** Schematic of Body Clock Measurement. Time-1 full model provides estimates of longitudinal BODN (at three-time points, green arrows). Time-2 full model provides estimates of BODN at time 2 and 3 (blue arrows). The orange arrow shows change in Body Clock. The red arrows depict the effect of incorporation of future information on a new disease on the Body Clock.

**Table 1:** Body Organs and The Corresponding Disease Levels Definitions Based on Disease Severity or Stage

| **Cardiovascular System (CVS)** |  |
| --- | --- |
| Hypertension<br>(HTN; 1-3) | 1: No HTN<br>2: SBP>140; DBP>90 or documented history of HTN and no medication<br>3: SBP>140; DBP>90 or documented history of HTN and medication<br>HTN: medication (hydropyridines calcium channel blocker (CCB), diuretics, angiotensin converting enzyme inhibitors, beta blockers, and other antihypertensive drugs) |
| Ischemic Heart<br>Disease (IHD; 1-3) | 1: No IHD<br>2: History of prolonged angina pectoris, angioplasty, or coronary artery bypass surgery (CABG)<br>3: Myocardial infarction (significant q wave on EKG, documented history of myocardial infarction, or nitrate medication) |
| Congestive Heart<br>Failure (CHF; 1-3) | 1: No CHF<br>2: CHF with preserved ejection fraction (EF)<br>3: EF < 50 or history of heart failure and use of digitalis, non-hydropyridines CCB |
| Arrhythmia<br>Based on<br>Electrocardiogram,<br>(EKG; 1-6) | 1: No arrhythmia<br>2: Sinus bradycardia<br>3: Elongated QTC (> 440 ms in men or > 460 ms in women)<br>4: Elongated P-R interval >200 m-second<br>5: Supraventricular, ventricular, or paroxysmal atrial tachyarrhythmia<br>6: Atrial fibrillation |
| Peripheral Artery<br>Disease (PAD; 1-2) | 1: No PAD<br>2: Left or right ankle brachial index (ABI) < 0.9 |
| **Cerebrovascular Accident (CVA)** |  |
| Stroke (1-4) | 1: No stroke<br>2: TIA with no history of severe stroke<br>3: History of documented stroke, elongated sudden numbness, or loss of speech with no paralysis or limb weakness<br>4: Paralysis or paresis in either limbs or any sudden and long lasting of motor neuron dysfunction such as cranial nerve (CN) CN-III, CN-IV, CN-VI (ocular movements, nystagmus and ocular convergence); CN-IX (Oropharynx symmetry); CN-VII (Facial asymmetry) |
| **Renal (Ren)** |  |
| Chronic Kidney Disease (CKD; 1-5)<br>Using CKD-EPI formula | 1: Estimated glomerular filtration rate (eGFR) > 90 (no CKD)<br>2: Stage-1: $60 \leq \text{eGFR} \leq 90$<br>3: Stage-2: $45 \leq \text{eGFR} \leq 59$<br>4: Stage-3: $30 \leq \text{eGFR} \leq 44$<br>5: Stage-4: $\text{eGFR} \leq 29$ |
| **Metabolic (Me)** |  |
| Diabetes Mellitus (DM;1-6) | 1: No DM<br>2: Impaired glucose tolerance test ($140 \leq$ at $\leq 2$ hr postprandial blood sugar $\leq 199$ (mg/dL))<br>3: DM with no complications<br>4: DM with CKD no retinopathy<br>5: DM with retinopathy no CKD<br>6: DM with both CKD and retinopathy |
| Hyperlipidemia (1-3) | 1: No hyperlipidemia |
|  | 2: Triglyceride (TG) >150 mg/dL, low density cholesterol (LDL) > 130 mg/dL, high density cholesterol (HDL) < 40 mg/dL for men and < 50 mg/dL for women, and no medication<br>3: TG > 150 mg/dL, LDL > 130 mg/dL, HDL < 40 mg/dL for men and < 50 mg/dL for women, and medication |
| **Gastrointestinal and Liver (GL)** |  |
| Liver (1-3) | 1: No liver disease<br>2: History of hepatitis or increased liver function tests 2 times more than normal range<br>3: Liver fibrosis (based on FIB4 scoring system)<sup>54</sup><br>4: Liver cirrhosis |
| Gastrointestinal disease (GID; 1-3) | 1: No GID<br>2: Gastritis or gastroesophageal reflux disorder (GERD)<br>3: History of peptic ulcer, current ulcer, or use of antacid medication |
| **Respiratory (Res)** |  |
| Chronic Obstructive Pulmonary Disease (chronic bronchitis or emphysema) (COPD; 1-3) | 1: No COPD<br>2: Wheeze with physical exam (Yale criteria) and no medication use<br>3: Wheeze with physical exam (Yale criteria) and use of medication (adrenergic drugs, leukotriene receptor antagonists, or others) |
| Adult-Onset Asthma (1-2) | 1: No asthma<br>2: Documented adult onset, still having asthma |
| **Dysthyroidism (Th)** |  |
| Hypothyroidism<sup>55</sup><br>(1-4) | 1: Euthyroid<br>2: Subclinical hypothyroidism: $4.5 \leq \text{thyroid stimulating hormone (TSH)} \leq 6.9$<br>3: $7.0 \leq \text{TSH} \leq 9.9$<br>4: $\text{TSH} \geq 10$ or use of levothyroxine |
| Hyperthyroidism<br>(1-2) | 1: Euthyroid<br>2: $\text{TSH} < 0.3$ or use of thiouracil or other antithyroid medication |
| **Hematopoietic System (He)** |  |
| Anemia (1-4) | 1: No anemia<br>2: Mild anemia (women: $11.0 \leq \text{Hemoglobin (Hb)} < 12$; men: $11.0 \leq \text{Hb} < 13$)<br>3: Moderate anemia: $8.0 \leq \text{Hb} < 11$<br>4: Severe anemia $\text{Hb} < 8.0$ (rarely seen in population studies) |
| Thrombocytopenia<br>(1-2) | 1: Normal platelet count<br>2: Platelet count $< 150,000$ without cancer or liver disease states |
| **Oral Health (Periodontitis: Pe)** |  |
| Periodontitis (1-3) | 1: Normal<br>2: Gingivitis or periodontitis<br>3: Edentulous, $>50\%$ tooth loss due to periodontitis |
| **Musculoskeletal System (MS)** |  |
| Osteoarthritis (OA) | 1: No OA<br>2: Pain more than 1 month or stiffness but not both, at either knee or hip joints<br>3: Both pain and stiffness at the knee or hip joints |
| Osteoporosis<br>(based on bone mineral density (BMD), total BMD) | 1: Normal bone mineral density<br>2: Osteopenia (BMD score $< -1.0$)<br>3: Osteoporosis (BMD score $< -2.5$) |
| at either vertebrae<br>L1-4, femoral neck,<br>or trochanter; 1-3) |  |
| **Sensory System (Se)** |  |
| Hearing (1- 4) | 1: Normal hearing<br>2: Decreased hearing ability from poor to deaf without a hearing aid<br>3: Normal to fair hearing ability with a hearing aid<br>4: Poor hearing with a hearing aid |
| Eye (1-5) | 1: Normal eye function<br>2: Cataract but no glaucoma or macular degeneration<br>3: Glaucoma but no macular degeneration<br>4: Macular degeneration but no glaucoma<br>5: Glaucoma and macular degeneration |
| **Central Nervous System (CNS)** |  |
| Depression (1-3)<br>CES-D: Center for<br>Epidemiologic<br>Studies Depression<br>Scale (CES-D) | 1: No depression<br>2: CES-D scores $\geq 16$ but no use of antidepressants.<br>3: CES-D score $\geq 16$ and use of antidepressants |
| Parkinson's<br>Disease (PD; 1-3) | 1: No Parkinson's<br>2: Documented history of PD (Hoehn and Yahr scale), mild symptoms, stage $\leq 3$ or no medication<br>3: Stage 4 or 5 PD, moderate to severe symptoms or use of medication |
| Global Cognitive Impairment (CI; 1-3) | 1: Normal global cognition<br>2: Total mini mental status (TMMS) < 24<br>3: Documented Alzheimer's disease (combined medical examinations)<br>*Vascular dementia is part of cerebrovascular accident |
| **Cancer (Can)** |  |
| Lung, Breast, Prostate, Stomach, Pancreatic, Leukemia, Thyroid, Ovary, Squamous Cell Carcinoma, melanoma | 1: No cancer<br>2: At least one adult-onset cancer |
| **Late-life Outcomes** |  |
| Disability | instrumental activities of daily living (iADL) $\geq 2$ or activity of daily living (ADL) >1 |
| Geriatric Syndrome | Experienced two injurious falls in past month, urinary or bowel incontinence |
| Functional decline<br>SPPB < 9 | Short Battery of Physical Performance, which includes chair standing (1-4), walking speed (1-4), and balance (1-4) with total score 12 |
| Mortality | Mortality any time after data used in this analysis |

With the premise that each disease impacts body systems heterogeneously, we determined coefficient estimates of lagged disease(s) at two time points (time-1 and -2) predicting longitudinal BODN (Fig. 1b). For each disease, we quantified a “total disease beta coefficient estimate” as well as “disease level estimates” to predict longitudinal BODN (from one lagged time to last follow-up), applying multilevel ordinal regression in the Bayesian inference framework with individuals as a model level (model levels are different than disease levels) and reported 95% credible intervals (CI), counterparts to confidence intervals (Table 1). Avoiding possible crude categorization, we used “monotonic effect” in a Bayesian inference framework that considers the ordinal predictor as a continuous variable and provides a total coefficient estimate from the first to the last level of an ordinal predictor. Then it introduces ordinal cut-points after encountering the data and provides the proportion of each ordinal level so that level-specific estimates can be quantified. First, to assess how time predicts BODN, a model included only time as the period from the lagged disease to the last follow-up. To understand how c-age, single-disease, or single organ-system diseases incorporate into longitudinal BODN, we separately developed models at time-1 and time-2 for c-age, single disease, single organ-system diseases, multiple-systems diseases, and finally global disease levels burden from all systems.

We obtained the degree to which each disease and its levels incorporate into longitudinal BODN. We then assessed each model’s performance by quantifying model weight, which indicates model predictivity, and compared each model with c-age to disentangle the predictive role of c-age versus lagged single disease, single system, and multiple systems to predict longitudinal BODN. The model performance assessment (leave-one-out cross-validation) ^20^ and model weight comparisons (Bayesian Stacking weights) ^21^ are described in the methods. Using the post-analysis prediction function in the model including time-1 and -2 global disease burden levels, we predicted longitudinal BODN for each individual, termed *Body Clock*, then determined change in Body Clock over time.

### Single Disease and Diseases of Single System Heterogeneously Incorporate into Longitudinal BODN

All single diseases and single-organ system diseases with heterogeneous magnitudes of beta estimates, significantly incorporated into longitudinal BODN (multimorbidity) except hyperthyroidism. C-age in age-only model at time-1 and -2 was a strong predictor of BODN (time-1 age: b=0.20±0.01, 95% CI=0.19–0.23; time-2 age: b=0.24±0.01, 95% CI=0.21–0.26; Tables S2). Diseases of a single system, e.g., cardiovascular system (CVS), also heterogeneously and significantly predicted multimorbidity at both time-1 and -2 (Table S2).

### Preceding Diseases and Disease Level Burdens of Multiple System Heterogeneously Incorporate into Longitudinal BODN

We assessed the degree to which disease levels of all organ systems incorporate into longitudinal BODN, quantifying posterior coefficient estimates and 95% CI. Overall, all time-1 diseases heterogeneously incorporate into BODN. However, hyperthyroidism, peripheral artery disease (PAD), and Parkinson’s, with wide credible intervals (including 0), had larger uncertainty in predicting BODN (Fig. 2a,b, Table S2a). The CVS had the largest frequency as a single organ system and was the main component of BODN at all age groups (Fig. S1, S2). However, there was variability in its disease estimates with hypertension (HTN, b=0.49, 95% CI=0.38–0.59) and congestive heart failure (CHF, b=0.32, 95% CI=0.20– 0.45) as the strongest predictors that incorporated into BODN.

**Figure 2.**
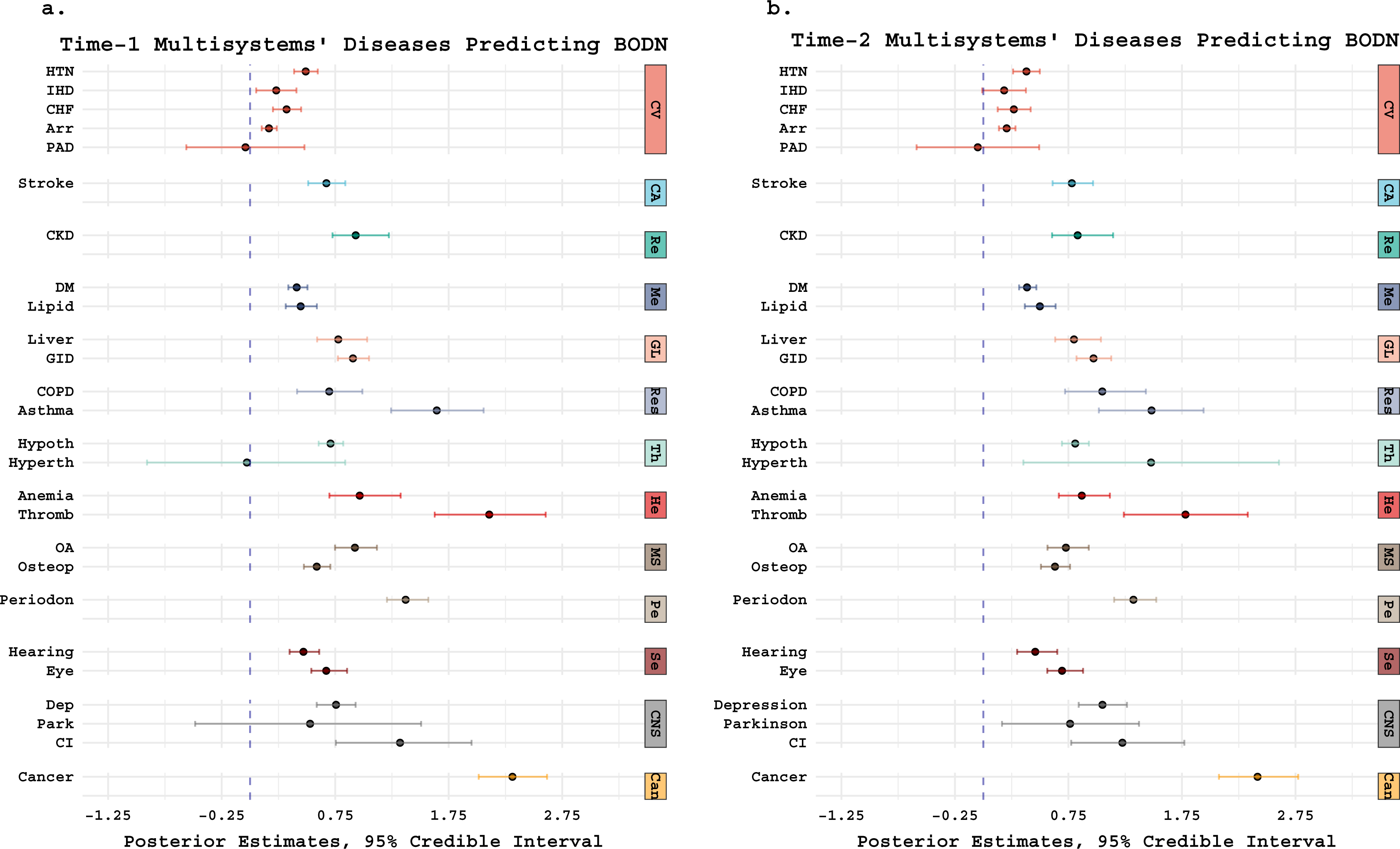
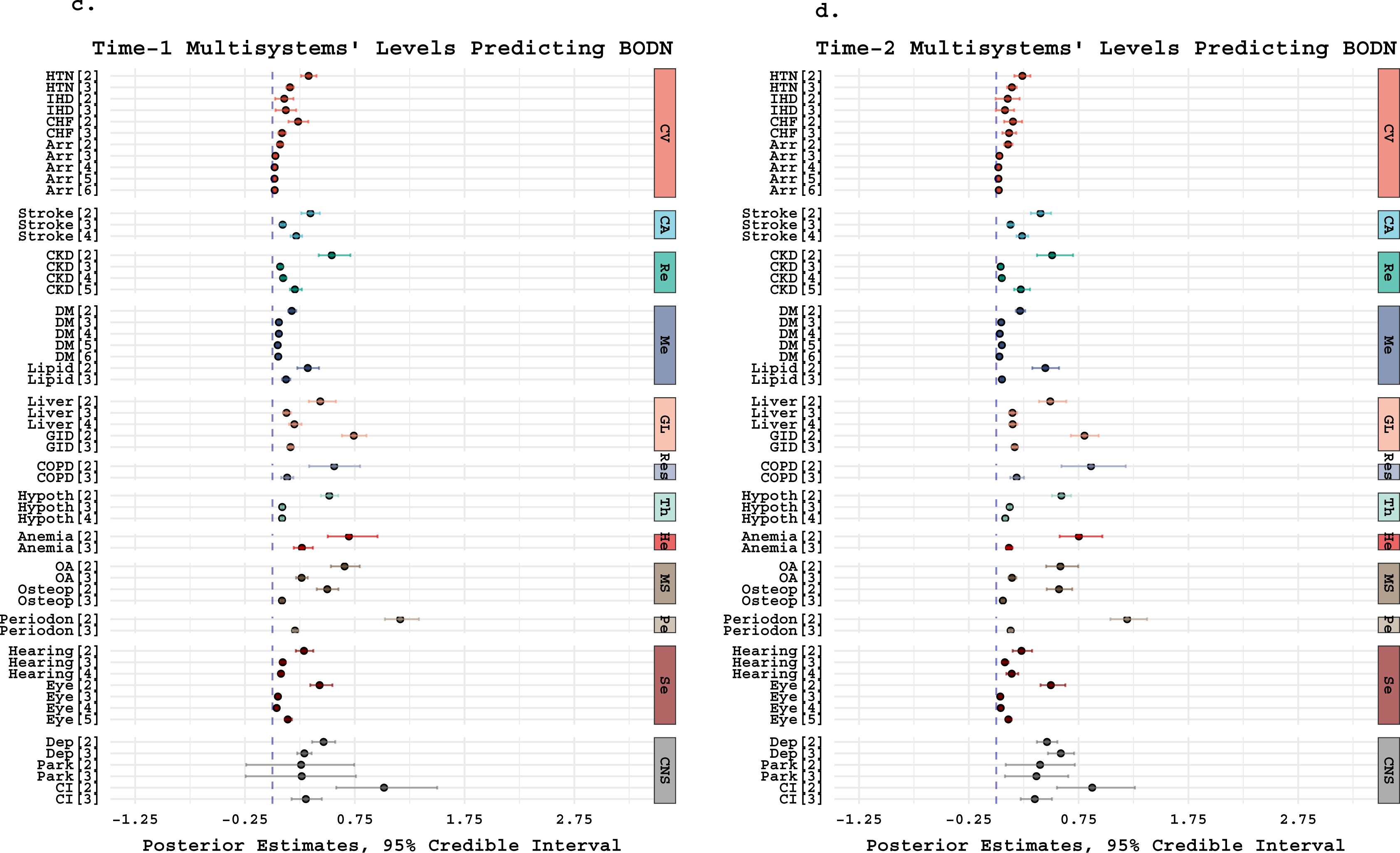
Lagged full models’ multisystem posterior estimates predicting longitudinal Body Organ Disease Number (BODN). The larger the posterior estimates with narrower 95% credible intervals (CI), the better the prediction is. (**a & b).** Total disease coefficient estimates, with 95% CI predicting BODN in the full models using Bayes approach at **a.** time-1 and **b.** time-2. Organ systems diseases have heterogeneous coefficient estimates predicting BODN. (**c & d).** Disease levels coefficient estimates heterogeneously incorporating into BODN at **(c)** time-1 and (**d)** time-2 full models. The significance is considered when 95% CI does not include 0. HTN: hypertension, IHD: ischemic heart disease, CHF: congestive heart failure, PAD: peripheral artery disease, Arr: arrhythmia, CKD: chronic kidney disease, DM: diabetes mellitus, GID: gastrointestinal disease, COPD: chronic obstructive pulmonary disease, Hypoth: hypothyroidism, Hyperth: Hyperthyroidism, OA: Osteoarthritis, Oteop: Osteoporosis, Periodon: periodontitis, Dep: Depression, Park: Parkinson’s disease, CI: cognitive impairment. Systems abbreviations: CV: cardiovascular system, CA: cerebrovascular accidents, Re: renal system, Me: metabolic, GL: gastrointestinal and liver, Res: respiratory, Th: dysthyroidism, He: hematopoietic, MS: musculoskeletal, Pe: periodontitis, Se: sensory, CNS: central nervous system, Can: Cancer.

Overall, milder states of organ system diseases (e.g., transitional ischemic attack, impaired glucose tolerance, mild anemia, subclinical hypothyroidism, cataract, mild liver disease, gingivitis without edentulous, osteopenia, and mild osteoarthritis, and poor hearing that improves with hearing aid) predicted multimorbidity with larger estimates than the more severe states.

Likewise, diseases with no treatment modalities had larger coefficient estimates predicting multimorbidity (BODN), as seen with HTN, type-2 diabetes mellitus (DM), hyperlipidemia, chronic bronchitis, gastrointestinal disease (GID), depression, and Parkinson’s (Fig. 2b, Table S2a). For example, HTN with no treatment (b=0.32, 95% CI=0.26–0.40) was more predictive of BODN than HTN with treatment (b=0.16, 95% CI=0.13–0.19; Fig. 2a, Table S2a,). Moreover, CHF with preserved ejection fraction (EF; b=0.23, 95% CI=0.14–0.32), common with aging, had a larger coefficient estimates than CHF with low EF (b=0.09, 95% CI=0.05-0.12). For arrhythmias, sinus bradycardia, followed by elongated QTc and atrial fibrillation, incorporated more significantly into multimorbidity than others.

Stage-1 age-related CKD, uncoupled from diabetic kidney failure, incorporated more strongly into multimorbidity (stage1; b=0.54, 95% CI=0.42–0.71) than stage-2 (b=0.07, 95% CI=0.05-0.09) and stage-3 (b= 0.10, 95%CI=0.08-0.13), followed by end-stage-renal disease (stage-4; b=0.20, 95% CI=0.16–0.27; Fig. 2a, b, Table S2a).

We detected a similar dynamic pattern in the time-2 full model integrating all diseases, but now hyperthyroidism (b=1.47, 95% CI=0.45–2.60) and Parkinson’s (b=0.76, 95% CI=0.16–1.37) incorporated significantly into multimorbidity (Fig. 2c,d, Table S2b). Model assessments showed that full models had the highest accuracy in predicting multimorbidity (Table S3, Fig. S3a-b).

Notably, while PAD strongly predicted multimorbidity as a single disease (time-1; b=3.68, 95% CI=2.52–4.81) or as part of a single organ system (time-1; b=1.36, 95% CI=0.27–2.43), its significance was eliminated in the full models (time-1: b=-0.04, 95% CI=-0.5–0.47; time-2: b=-0.04, 95% CI=-0.58–0.49). One could speculate that shared pathophysiology among one system’s diseases might change its significance when collapsed into one model. To test this, we used Bayesian stacking that finds model-specific weights. The model has zero weight if it is not adding more predictive information to the other models. Using Bayesian stacking weights for all cardiovascular-related single-disease models, we assessed their usefulness in predicting multimorbidity. The models’ weights for PAD and ischemic heart disease (IHD) turned to 0, but allocating weights to HTN, CHF, and arrhythmia by 49.7%, 48.3%, and 2%, respectively, suggesting shared pathophysiology between CVAS diseases.

### Age Outperforms Time, Single Disease, and Single Organ System to Predict Longitudinal BODN

Because most chronic diseases manifest at older ages, we hypothesized that c-age is a stronger predictor of longitudinal BODN than any single disease or single-system diseases.

Bayesian Stacking of model weights revealed that the age-only model outweighed any single disease or single-system diseases, suggesting that c-age is the driving force for health deterioration. Among all single diseases, eye, kidney, and periodontitis carried larger weights predicting BODN, followed by GID and DM (Fig. 3; Table S2). Comparing time and c-age model weights revealed c-age is a stronger predictor than time, at both time-1 and time-2 (Table S3).

**Figure 3.**
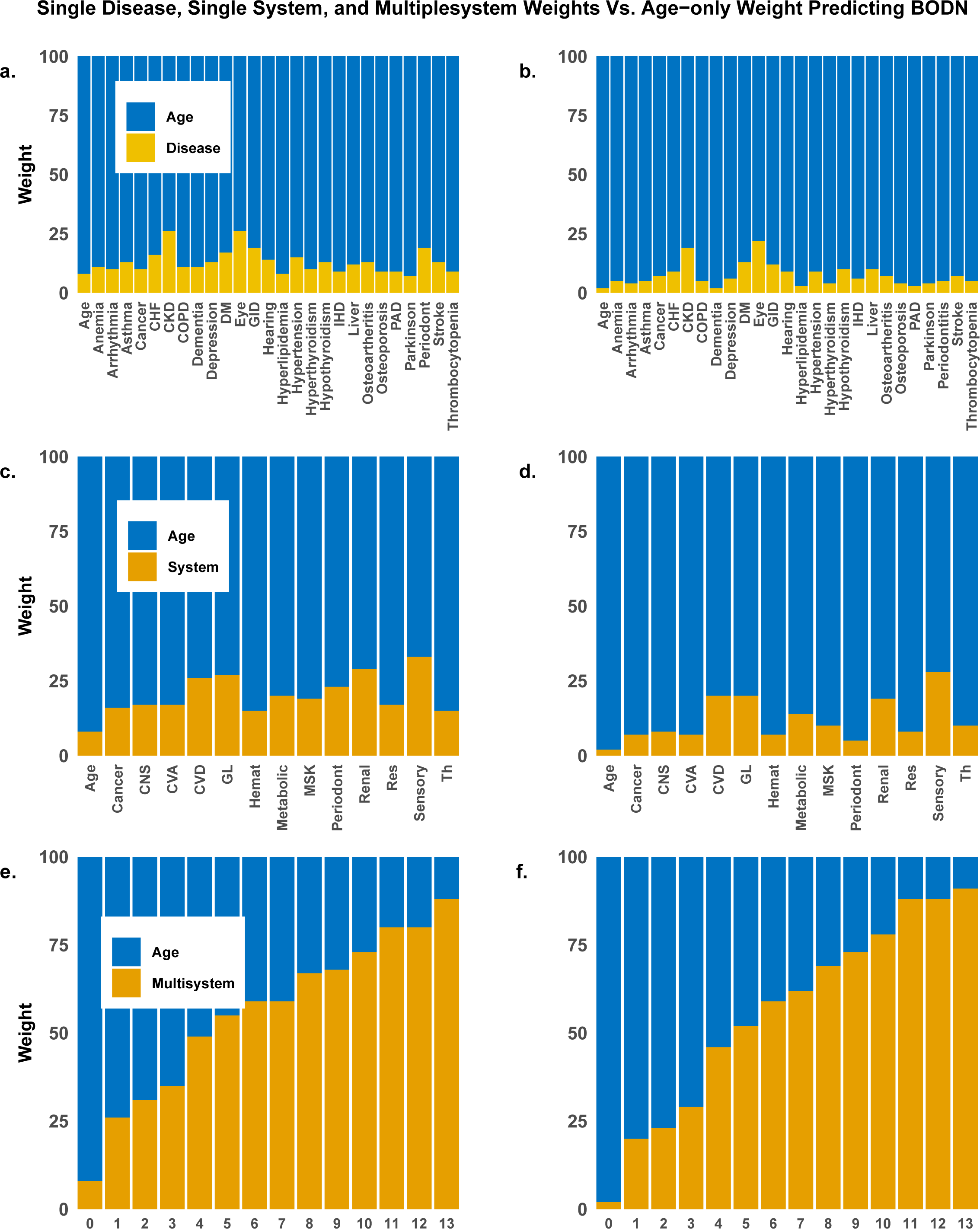
Bayesian Stacking weights for time-1 and time-2 models compared to the age-only model. (**a)** Time-1 model weights for every single disease compared to age-only model weight predicting BODN. (**b)** Similar comparisons for time-2 models. (**c)** Time-1 single-system model weight versus age-only model. (**d)** Similar comparisons for time-2 models. C-age weight outperformed single disease and single organ-system weights. In time-2 models, when people were older, the age-only model outweighed single disease even more than time-1 (younger age). € Time-1 stepwise multiple organ-system weights versus age-only model. (**f)** Similar comparisons for time-2 models. Time-2 full model outweighed c-age the most, indicating the global burden of organ-system diseases measure the rate of aging better than any single disease or single system. CHF: congestive heart failure, CKD: Chronic Kidney disease, COPD: Chronic Obstructive Pulmonary Disease, DM: diabetes mellitus, GID: gastrointestinal disease, IHD: ischemic heart disease, PAD: peripheral artery disease, CNS: central nervous system, CVA: cerebrovascular disease, CVD: cardiovascular disease, GL: gastrointestinal and Liver, Hemat: hematopoietic system, MSK: musculoskeletal system, Periodent: periodontitis, Res: respiratory system, Th: Dysthyroidism. The numbers in graphs e and f indicates the number of systems with morbidities in Body Organ Disease Number (BODN).

### Diseases of Multiple Systems Outperform C-Age in Predicting Longitudinal BODN

When we used one system’s integrated diseases, the models performed better than any single disease to predict multimorbidity. With the premise that diseases of one system can impact all other systems, we hypothesized that global burdens of disease levels from multiple systems would better predict future BODN than a single disease, single-system diseases, or c-age. In the stepwise analyses, multiple-system integrated diseases better predicted longitudinal BODN than any other single model shown by model performance and model weight compared to age-only models. Finally, integrating all lagged diseases, the full models outperformed c-age to predict longitudinal BODN (Figs. 3e,f, Table S2), suggesting that this is a global measure capturing the rate of biological aging and its effect on health. Likewise, global disease burdens at time-2 predicted BODN better than c-age so that the time-2 full model outweighed c-age in predicting BODN with a larger weight than the time-1 model (when participants are younger, and time is longer). Together, this shows that intrinsic biological age does increase with c-age and time, but it is steeper with advancing c-age (Figs. 3e,f). In-sample (simulated BLSA data) and out-of-sample validations (InCHIANTI data) confirmed these findings (Fig. S4).

### Body Clock in Men and Women and Change in Body Clock Over Time

We used burdens of disease levels at each time point to predict longitudinal BODN at the individual level, termed *Body Clock*. Therefore, Body Clock can be based on time-1 diseases or time-2 disease levels. The change in Body Clock is quantified based on both time-1 and time-2 predicted longitudinal BODN and its change from baseline (Fig 1b). In the multilevel analyses, individuals are included as a model level. The spaghetti plots depict the heterogeneity in individuals’ Body Clock’s trajectories even within the same c-age spectrum, revealing a unique rate of aging for each person (Fig. 4a,b). Quantifying a personalized Body Clock, from sub-clinical states to severe diseases, allows us to measure the process of aging at any age. Using quadratic spline regressing Body Clock over c-age revealed that Body Clock increases non-linearly with c-age over lifespan from the 40’s to the 90’s, in both women and men (purple overlay, Fig. 4a,b). The association of Body Clock based on time-1 diseases and c-age groups with 10-year intervals showed increase in Body Clock with c-age over the lifespan (men: b=0.95, 95% CI= 0.79-0.95; women: b=0.90, 95% CI=0.89-0.98). In addition to fixed effect of c-age on Body Clock, we assessed the random effects of each c-age, including age (years) as model level, to quantify variability of Body clock within the same c-age and between c-ages. The wide 95% credible interval depicts significant within-age variability of Body Clock (does not include 0) after age 67, increasing with c-age (Fig 4c).

**Figure 4.**
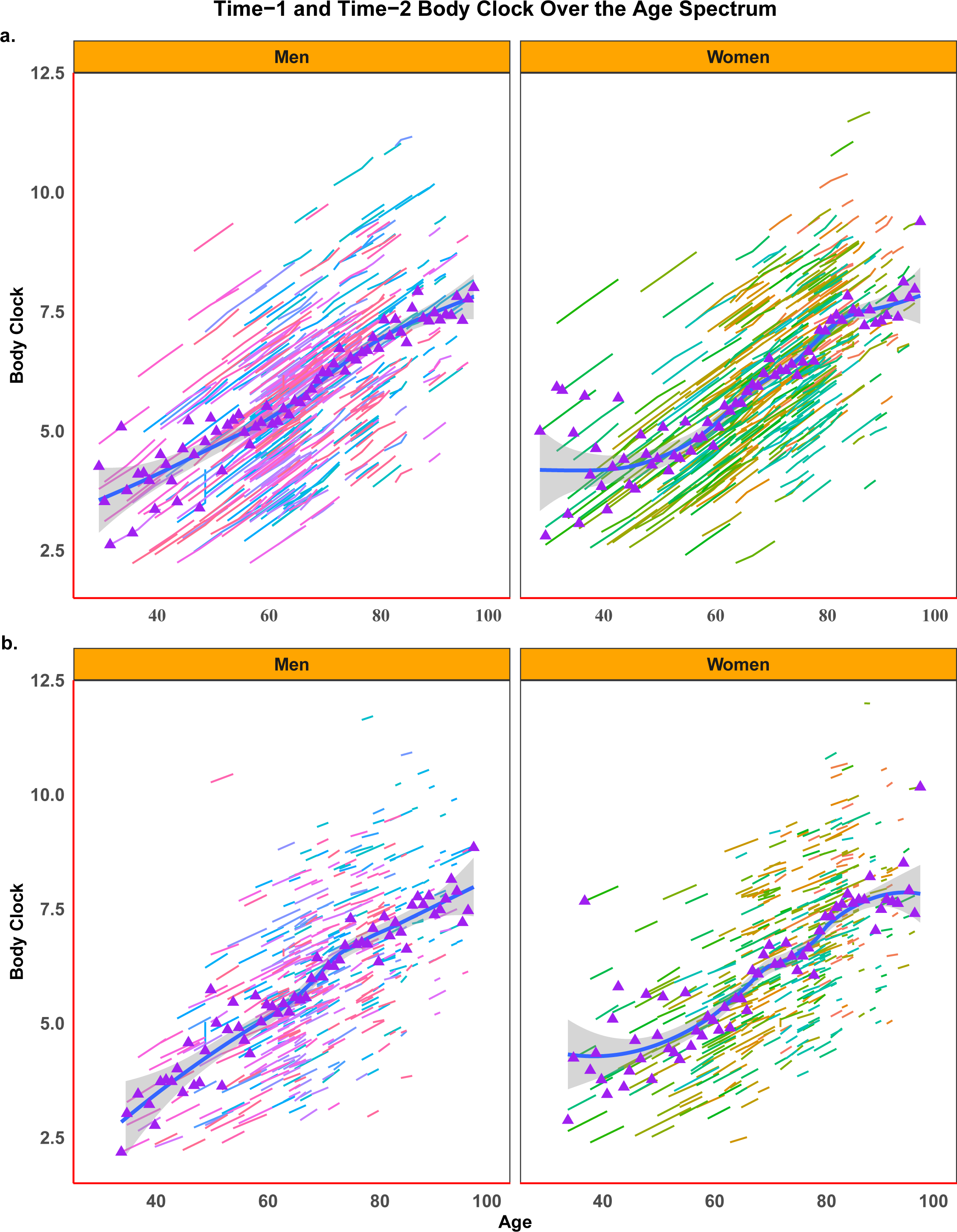
Body Clock and chronological age. (**a)** Body Clock using time-1 organ-system disease levels predicting longitudinal BODN. (**b)** Body Clock using time-2 organ-system disease levels predicting BODN. The spaghetti plots show individual-based integrated predicted estimates of BODN as sporadic lines (Body Clock). The quadratic spline regression line (purple overlay) with a 95% credible interval is a population-based Body Clock in men (left) and women (right). Body Clock increased with advancing age, even in younger age groups with substantial interindividual variability. **(c)** Random effect of age on change in the Body Clock. There are within the same age and between age variability in Body Clock. The random variability of the Body Clock within the same age is significant after age 67.

### Body Clock Predicts SPPB, Disability, Geriatric Syndrome, and Mortality

To assess whether Body Clock predicts late-life age-related outcomes, we performed multilevel cubic-spline Cox-regression hazard models within the Bayesian framework ^22^ to predict Short Battery of Physical Performance (SPPB) <9 ^23^; disability (Activities of Daily Living scale [ADL] >1 or Instrumental Activities of Daily Living scale [IADL]>1]^24^; geriatric syndrome (having injurious fall or urinary/bowel incontinence ^25^; and mortality (Fig. 5). The results showed that people with higher Body Clock are at higher risk of SPPB<9 (hazard ratio [HR]=2.30, 95% CI=1.85–2.90, SD=18.0); geriatric syndrome (HR=1.70, 95% CI=1:55–1.80, SD=3.30); disability (HR=1.8, 95% CI=1.44–1.80); and mortality (HR=1.71, 95% CI=1.42–2.11, SD=1.68), showing Body Clock’s capability as an initial measure of health to predict late-life outcomes. To evaluate a dynamic reciprocal relation between Body Clock and SPPB, we examined the association between time-1 SPPB<9 with the change in Body Clock, revealing that lagged functional impairment also predicts change in Body Clock (SPPB<9) (b=0.96, 95% CI=0.57-1.35).

**Figure 5.**
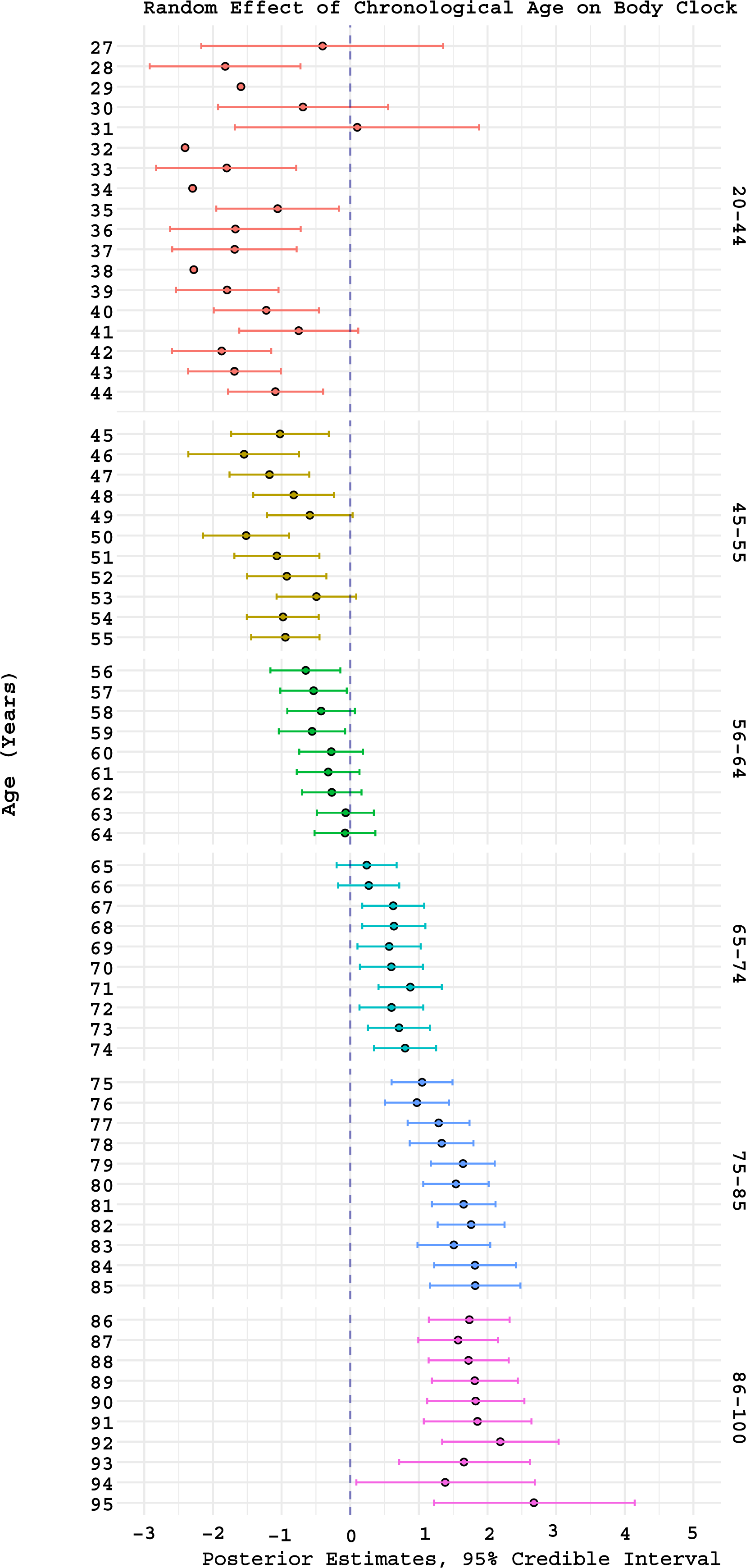
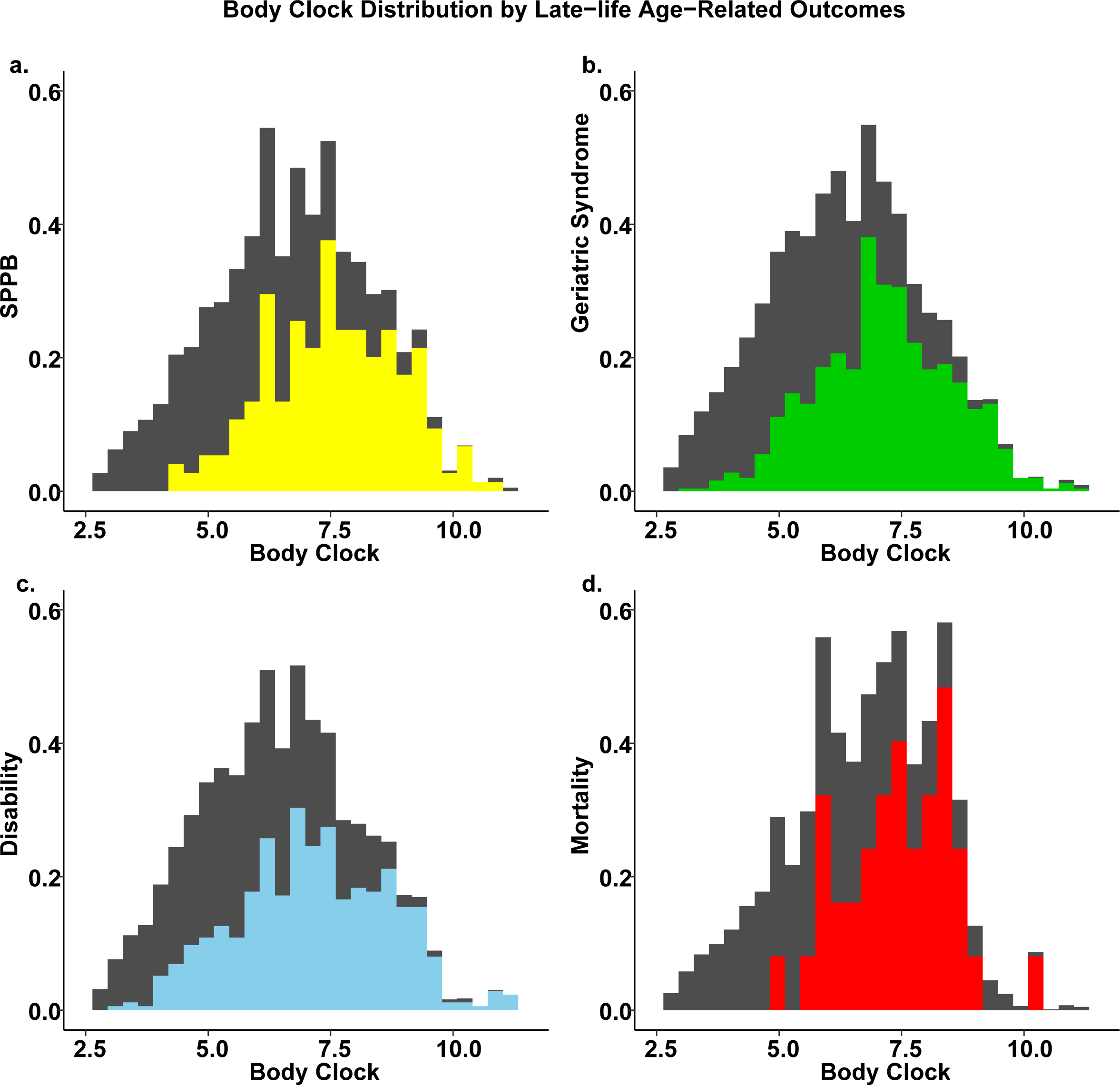
Density Distribution of Estimated Body Clock by Mortality, Geriatric Syndrome, Disability, and Functional Decline (SPPB < 9). The distribution of Body Clock with a. Short Battery of Physical Performance (SPPB) < 9, b. Geriatric Syndrome, c. disability (Activity of daily living (ADL) or instrumental activity of daily living (iADL), and d. mortality as binary outcomes using the Cox model in the Bayes Framework. An average Body Clock of more than 7.0 is highly significant in all models. However, some individuals have the resilience to developing these late-life outcomes even at larger Body Clock values. In all graphs, the gray area is the normal state of the corresponding late-life outcome.

## Discussion

We applied Bayesian inference to longitudinal data with well-defined clinical information and developed a validated personalized tool that matches multimorbidity with fast aging.

Approaching multimorbidity as a global organ system health outcome, we defined body organ disease number (BODN) and quantified the degree to which each disease levels incorporate into longitudinal BODN. Using the post-analysis prediction function, we defined individual-based predicted BODN, termed Body Clock. The heterogeneity in disease estimates and the interindividual variability in Body Clock supports the idea of heterogeneity in rates of both organ and individual aging. C-age was a stronger predictor of BODN compared to any single disease or single organ system. However, global burden of disease levels and their levels outperformed c-age, allowing to predict individual-based BODN and quantify Body Clock as a rate of biological aging uncoupled from c-age, which predicts late-life outcomes.

The Charlson Index ^26^, a common hospital-based multimorbidity index, includes c-age weighted diseases based on a hazard ratio for predicting 1-year mortality. The category-allocated weighted value might bias the multimorbidity score by selective-mortality, disregarding early-onset changes at younger ages. However, Body Clock, uncoupled from c-age, reflects progressive health impairments at any age independent of mortality. The Cumulative Illness Rating Scale for Geriatric (CIRS-G) is another hospital-based index using medical chart reviews of 13 systems. It includs both acute and chronic disease burdens, to calculate a total score, a severity score as an average over the severity of the diseases (0 to 4), and a comorbidity index as mean severity score of 2 or more. CIRS-G can skew the severity scores toward the acute condition that originally caused the hospitalization. For example, current acute myocardial infarction is scored 4 while congestive heart failure, an age-related health deteriorating chronic disease is scored 2.

Moreover, averaging over the severity of the diseases assumes that the disease severity levels are equidistant. However, we showed substantial heterogeneity in disease levels’ estimates that predict body organ systems. Moreover, averaging the disease severity levels might bias the comorbidity index by underestimating chronic diseases relative to acute morbidities. Disability, a late-life outcome, was also included as part of the disease severity score, while we predicted BODN as Body Clock independent of late-life outcomes ^27^. Nevertheless, our suggestive algorithm could be applied to patients’ chart review at hospital or clinic to predict late-life outcomes.

There is consensus that functional integrity without disability is one key component of healthspan ^28, 29^. Hence, some multimorbidity tools, such as the multimorbidity-weighted index, employed physical function to weight diseases to measure multimorbidity ^9, 30^. However, one might have physical function resilience when compensatory physiologic strategies in some organs cope with the effects of damage accumulation in other organ systems ^6^, leading multimorbidity-weighted estimates biased by physical functional states. Moreover, weighting organs by physical function status skews the metric toward more irreversible late-life health states. Notably, counterintuitive finding from a clinical trial, the LIFE trial, aimed at targeting late-life functional decline with structured physical activity intervention, reported paradoxical results for multimorbidity with further hospitalization and mortality in the group with higher multimorbidity burdens ^31^. However, we showed that declining physical function predicts increased Body Clock, reinforcing its bidirectionality ^32^, suggesting that age-related impairments should be measured and intervention(s) initiated before irreversible declines occur. Therefore, we developed Body Clock independent of physical functioning, yet it predicted late-life outcomes with high accuracy.

Some indices use diseases that predict in-hospital mortality as multimorbidity measures. Any health tool based on mortality ^10, 12, 33^ is more life-span oriented and can bias multimorbidity measures in survivors of potentially fatal diseases who have more extended longevity. Moreover, the mortality rate itself can be confounded by the type of hospital facilities, type of care, individual choices for care and technological advances over time. As such, we quantified Body Clock based on a prediction of future BODN as a measure of systems multimorbidity rather than predicting mortality. Other indices apply summation of the number of diseases while disregarding the possible heterogeneity in the degree to which these diseases incorporate into multimorbidity ^8^. We showed such heterogeneity exists for chronic diseases and their levels, accentuating the variability in organ aging. For example, the cardiovascular system (CVS) with the highest system morbidity frequency across all age spectrum includes five diseases. However, CHF with preserved ejection fraction, mainly reported in older adults ^34^; HTN without treatment; age-related arrhythmias such as sinus arrhythmia; and atrial fibrillation ^35, 36^ were the levels that predicted BODN most strongly, indicating that merely summing the number of diseases can over or underestimate multimorbidity.

Alternatively, several tools have been developed to reveal the rate of aging, mainly employing biochemical and/or physiological biomarkers, which might not disentangle damage from compensatory mechanisms ^12, 13, 15^. Moreover, biomarker-based tools can fluctuate due to transient stress levels and, therefore, might not distinguish between immediate compensatory mechanisms and longer-term health perturbations. One could speculate that those who do not respond well to stress and have lower values for these markers might have worse health problems.

Data showed that diseases at sub-clinical states or without treatment have larger estimates for predicting multimorbidity. Notably, clinical trials have shown that mild hypertension strongly associates with cognitive impairments ^37^ or suggested that intensive HTN treatment is protective for cognitive impairments and kidney function ^38–40^. Our results also endorse the idea that sustained exposure to subclinical disease levels might result in chronic wear and tear causing damage accumulation, and culminates in variable impairment in some organ systems, manifested as heterogeneity in organ disease levels’ estimates, and change in personalized Body Clock.

Moreover, the sub-clinical states with large estimate magnitudes and wide uncertainty at younger ages suggest that some organ systems might maintain functional capacity to respond to accumulated damage in other organ systems with diminished resilience. Ferrucci’s team has shown that the resting metabolic rate (RMR) at disease-onset increases but the RMR declines with advancing chronic diseases over time with possible exhaustion in organ function at subcellular levels ^41^. Likewise, heterogeneity in disease-level estimate magnitudes suggests that at older c-age, the damaged state may reach a plateau with perturbed organs’ reserve to function and no repair capacity remains to respond to further damage. Age-related changes at sub-cellular and tissue levels can be manifested as physiological changes and finally as irreversible pathology accumulation. Therefore, it is not unrealistic to claim that Body Clock, an integrated estimates of disease levels predicting BODN, can capture allostatic overload, which is dramatic, cumulative physiological dysregulation, outstripping the convergence of stress and adaptation ^6^. Therefore, additional body stressors, such as diseases of other organs, medicines, infection, and other socio-environmental stressors, can be manifested as unique rate of increases in Body Clock.

For the first time, Body Clock matches multimorbidity with fast aging at individual level. Applying multilevel Bayesian inference to medically and clinically well-defined phenotypes with cross-population consistency showed that integrated disease levels of body organ systems outperform c-age to predict future BODN. It also unraveled heterogeneity in organ systems aging, endorsing multimorbidity as a clinical manifestation of the rate of intrinsic aging that was otherwise hidden with a single disease or a single organ-system approach. Furthermore, as an individual-based metric of damage accumulation, Body Clock can be applied to distinguish biomarkers of damage from those of resilience.

There are limitations to this study. One is that the time between follow-up points were not equidistant. However, by including time in the model and calculating personalize-based estimates we still could quantify Body Clock with high accuracy. Also, the BLSA is a study of healthy aging, recruiting initially healthy individuals. Hence, the general population with more severe conditions and hereditary diseases might have more accelerated Body Clock over c-age or time. However, because we quantify Body Clock at individual level, Body Clock can be updated whenever new personalized information on new disease levels is available.

Body Clock can be used to further disentangle variability in biological aging and its underlying mechanisms, including genetics and environmental factors, and creating new individual-based, multifaceted biomarkers and tailored interventions and can be updated and used as a potential assessment tool for geriatric clinics, hospitals, and clinical trials in a precision medicine approach.

## Material and Method

### Study Population

We used information on physical examinations, laboratory tests, and medical records collected for the participants of the Baltimore Longitudinal Study on Aging (BLSA), a cohort study of healthy aging in community-dwelling participants^29^. For external validation, we used the Invecchiare in Chianti (InCHIANTI) aging study, a prospective population-based study of older persons in Tuscany, Italy (as reported previously)^42^. Study participants have provided consent form. The data are available through BLSA and InCHIANTI website applications.

The BLSA study recruited individuals ≥20 years who were followed quadrennially if younger than 60, biennially if 60–79, and yearly if ≥80 or who developed the disease at any age. Our analyses included 456 men and 451 women (*N*=907) with three available subsequent visits with dynamic intervals (mean 3.9 ± SD 2.9 years) range (1-13) years of follow-ups. Mortality follow-up continued for 3.1 ± 2.3, range 1-9 years after the last visit through 2019.

### Body Organ Disease Number (BODN)

We defined disease states for 11 organ systems, two distinct diseases, cerebrovascular accidents (CVA) and adult-onset cancer. CVA shares pathophysiology and symptoms between CVS and CNS, so to avoid any misclassification and as a distinct entity, it was counted as one system. With distinct mechanisms from other age-related processes, cancer of any organ was also considered separate from other organ system diseases.

The organ system diseases and the levels of diseases are summarized in Table 1.

BODN is determined as the number of organs with at least one impairment as a new measure of multimorbidity. Pathology at the specific organ level was established based on predefined impairment criteria aligned to those that either deviates from normal at a younger age or is used for disease diagnoses in clinical practice. A specific organ system contributed to BODN, a global health index if it had at least one positive criterion. We then assigned organ-specific diseases an ordinal number for a level or stage of disease (e.g., no hypertension, hypertension with no treatment, and hypertension with treatment would be coded 1, 2, and 3, respectively; Table 1).

### Statistical Analyses

#### Bayesian inference

We used a Bayesian approach in developing our new multimorbidity tool. We used BODN as an ordinal outcome and time-1 or time-2 disease levels as lagged ordinal predictors, adjusting for the time but excluding c-age. We used multilevel ordinal regression and individuals as model level in a Bayesian framework to develop various models ^43^, assessed the models’ predictive accuracy using leave-one-out cross-validation (LOO-CV) ^20^, quantified model weights, and compared all models with c-age and the time. Including all disease levels in one model, we obtained post-analyses predicted BODN at individual levels to quantify individual-based Body Clock.

We evaluated the models using in-sample (using the BLSA parameters to predict BODN in simulated data) and out-of-sample accuracy (using BLSA parameters to predict BODN in InCHIANTI data) with “predictive checks,” as part of the usual Bayesian workflow ^44, 45^. We then determined the model-specific weights, performed model comparisons using the Bayesian stacking approach, and compared all models’ weights with the age-only model described in the supplementary information (SI). We used the Bayesian Cox-proportional hazard model to predict late-life age outcomes.

#### Bayesian Inference

A Bayesian model comprises two components: 1) Prior knowledge on the estimates (parameters), as the information before observing the data P(ϴ) where ϴ denotes the parameters; and 2) the likelihood P(Y|ϴ) of the information contained in the data (Y). Using the Bayes formula, we can obtain the posterior distribution of the parameters P(ϴ|Y), which can be updated when encountering new data. Moreover, the Bayes approach provides a distribution of estimates rather than a point estimate, and uncertainty around the estimates known as the credible interval (CI).^46^

### Bayes Approach for Multilevel Ordinal Outcome and Ordinal Predictors

With ordinal outcomes, the difference between the orders might not be equidistance. In other words, moving from having one organ with at least one disease to the next organ with disease is not necessarily equal to moving from having two or three organ morbidities to the next organ morbidity. We used a Bayesian ordinal model that assumes the observed ordinal outcome Y (i.e., BODN) is derived from categorizing a latent continuous variable ^Ỹ^, here BODÑ ^43^. Therefore, we used longitudinal, multilevel ordinal regression models with BODN as an ordinal outcome and each person as a level using “brms” Bayesian software package, built on top of Stan, a probabilistic programming framework for Bayesian inference ^45, 47–49^. Also, considering that the levels of ordinal predictors might not be equidistant, we used a function called “monotonic effect” implemented in the Bayesian “brms” software package published previously ^47, 50^, which assume that the ordinal predictor is a latent continuous variable with a posterior estimate (total beta coefficient for the ordinal variable). When encountering information from the data, a cut-point is introduced using the “simplex function” ζ, so that ζ *i* ∈ [0,1]. Therefore, ζ is the posterior probability of each level, and *i* is the number of ordinal categories. We can quantify the posterior estimate (beta coefficient) of each disease level through multiplying the total posterior estimate by the proportion of each level ζ*i*, e.g., HTN has three levels, the first level is the reference [normal]; the second and the third level proportions are ***ζ***% and 1-***ζ***%, respectively. With the posterior estimate ϴ (total beta coefficient) of the latent continuous HTN, the posterior estimate of each HTN level would be (ϴ * ***ζ***%) and (ϴ * 1-***ζ***%), respectively^50^. Except peripheral artery disease (PAD), hyperthyroidism, cancer, thrombocytopenia, and asthma, which are binary, all other diseases are ordinal (Table 1).

Using all disease-level burdens, we obtained predicted BODN for each individual as a level, termed *Body Clock* at each time-point and change in Body Clock over time. We used change in Body Clock to predict late-life outcomes such as Short Battery of Physical Performance (SBPP), geriatric syndrome, disability and mortality, using multilevel Cox-Hazard Models in Bayesian inefrence framework ^22^.

### Prior Knowledge of Predictive Estimates of BODN

We determined the prior distribution for each parameter, including intercept and beta coefficients using weak priors. For the beta coefficients and intercepts classes, the prior estimate with a normal distribution (mean: μ = 0, variance σ = 10), and for the class standard deviation (sd) related to varying subjects, the half-Cauchy (0,10) were used. The uniform prior with a Dirichlet distribution was used for ordinal predictors (i.e., (2, 2) for ζ1 and ζ2 [simplex parameters] for a three-level predictor).

We used a dynamic Hamiltonian Markov chain Monte Carlo (MCMC) algorithm ^48, 49^ to obtain posterior draws. We performed the analyses with a minimum of five chains and a minimum of 5,000 iterations. We obtained mean posterior values from the posterior distribution of the parameter estimate ϴ if the distributions were normal but reported median if not normal.

### Model Inference Diagnostics

To assess the computational success and resulting inferences, we evaluated the convergence diagnostic Rhat and the effective sample size measures for each estimated parameter ^51^.

### Model Comparisons

Each Bayesian model has its own predictive performance on a logarithmic scale called estimated log predictive density (ELPD). The ELPD estimates the predictive performance of future data, and its standard error (SE) quantifies the uncertainty in knowing the future BODN exactly. Leave-one-out cross-validation (LOO-CV)^52^, a variant where each observation takes the role of the validation set in turn and leads to a natural single prediction approach, was used for each model evaluation and also for model comparisons. As MCMC is time-consuming, Pareto smoothed importance sampling (PSIS) was used for faster LOO-CV estimates ^53^. If the PSIS diagnostic indicated an unreliable PSIS estimate (k>0.7), we used MCMC for a slower but more accurate computation. To compare each model with the age-only models, we used LOO-CV and comparison of models’ ELPDs to obtain ELPD_DIFF, which shows ELPD differences between each model and the age-only models. The models with larger ELPD are optimal, and a negative ELPD_DIFF favors the first model in the comparison.

### Model Evaluation and In-Sample and Out-of-Sample Predictive Checks

To assess the robustness of the model to predict BODN, we used the time-1 full model based on BLSA data to predict BODN in the InCHIANTI data as out-of-sample validation. We compared ELPDs of the models using LOO-CV. Additionally, we used “posterior predictive checking”^44^ implemented in “brms” software package to compare the posterior predictive density of simulated data to density estimates of the observed data (in-sample evaluation of BLSA) or predict new data (InCHIANTI data; out-of-sample validation).

### Bayesian Stacking Weights to Compare Models

Bayesian stacking is useful for computing model weights for several models simultaneously by optimizing the estimated LOO-CV predictive performance of the weighted model combinations.

Here we computed stacking weights for each kind of model paired with their corresponding age-only models (single-disease, single-system, stepwise multisystem, and full models separately).

The Bayesian stacking approach provides the model-specific weights versus the age-only models and informs the models with the best predictive performance.

## Data Availability

The data are available upon formal request to BLSA and InCHIANTI studies.

## Acknowledgements

We appreciate BLSA and InCHIANTI study participants and investigators and staff. We acknowledge Dr. Stefania Bandinelli for InCHIANTI study data and Elisa Fabbri for her previous work on multimorbidity in BLSA and InCHIANTI. ShS Acknowledge Intramural Research program of National Institute of Aging for supporting her Special Volunteer research activities.

Shabnam Salimi is funded by NIA-B K01AG059898.

This research was supported in part by Intramural Research program of the National Institute of Health, National Institute on Aging.

## Author Contributions

ShS developed the concept of Body Clock as fast rate of aging, chose the statistical approach and model selection, did data management, analyzed the data, designed the graphs, and wrote the main body of the manuscript. VA: supervised the analyses in Bayesian inference, advised on the methods, and communicating the statistical analyses. SM: contributed to the multimorbidity concepts, interpretation of the results, discussion, writing, and editing the manuscript. FL: Contributed to the multimorbidity concept as the rate of aging, interpreting results, writing, and editing the manuscript.

## Competing Interests Statement

There are no disclosures to declare. This material should not be interpreted as representing the viewpoint of the US Department of Health and Human Services, the National Institutes of Health, or its represented agencies.

## Supplemental Materials

### Results

Table S1 depicts the descriptive baseleine characteristics of Baltimore Longitudinal Study of Aging by baseline age groups.

**Table S1.** Baseline Characteristics of Baltimore Longitudinal Study of Aging

|  | **20-44**<br>**(n=68)** | **45-55**<br>**(n=137)** | **56-64**<br>**(n=230)** | **65-74**<br>**(n=278)** | **75-85**<br>**(n=143)** | **>85**<br>**(n=51)** |
| --- | --- | --- | --- | --- | --- | --- |
| Follow-up (years) | 5.8±4.1 | 5.4±3.7 | 4.1±2.7 | 3.5±2.2 | 5.0±0.37 | 5.2±0.7 |
| Mean ± SD<br>(range) | (1-15) | (1-15) | (1-14) | (1-14) | (1-9) | (1-11) |
| Sex |  |  |  |  |  |  |
| Female % | 51.5 | 39.4 | 38.7 | 56.8 | 62.2 | 51.0 |
| **CVD** |  |  |  |  |  |  |
| *HTN* | 19.1 | 27.0 | 44.8 | 54.3 | 59.5 | 62.7 |
| *IHD* | 0 | 2.9 | 10.4 | 14.4 | 28.7 | 25.5 |
| *CHF* | 36.8 | 40.2 | 50.4 | 51.8 | 60.2 | 80.4 |
| *Arrhythmia* | 27.9 | 24.8 | 33.0 | 42.8 | 74.5 | 49.0 |
| *PAD* | 2.9 | 1.46 | 0.87 | 2.9 | 9.1 | 13.7 |
| **CVA** | 4.4 | 4.4 | 4.3 | 9.4 | 17.5 | 17.6 |
| **CKD** | 32.4 | 66.4 | 73.1 | 92.8 | 100 | 96.1 |
| **Metabolic** |  |  |  |  |  |  |
| *DM* | 17.6 | 26.3 | 39.1 | 43.2 | 48.3 | 53.1 |
| *Hyperlipidemia* | 35.3 | 28.5 | 33.5 | 28.0 | 21.85 | 29.4 |
| **GL** |  |  |  |  |  |  |
| *Liver* | 2.9 | 11.7 | 13.9 | 16.6 | 32.9 | 15.7 |
| *GID* | 4.4 | 16.8 | 20.0 | 21.6 | 25.2 | 15.7 |
| **Respiratory** |  |  |  |  |  |  |
| *COPD* | 0 | 2.2 | 1.6 | 4.7 | 9.1 | 11.8 |
| *Adult Asthma* | 8.8 | 11.0 | 6.5 | 4.7 | 7.0 | 3.9 |
| **Dysthyroidism** |  |  |  |  |  |  |
| *Hypothyroidism* | 5.8 | 12.4 | 14.8 | 15.5 | 24.5 | 25.5 |
| *Hyperthyroidism* | 0 | 2.9 | 0 | 0.7 | 2.1 | 3.9 |
| **Hematopoietic Diseases** |  |  |  |  |  |  |
| *Anemia* | 10.3 | 8.0 | 6.0 | 8.6 | 15.4 | 23.0 |
| *Thrombocytopenia* | 5.6 | 2.9 | 3.5 | 5.0 | 6.3 | 3.9 |
| **Oral Health** | 20.6 | 22.6 | 34.8 | 35.3 | 34.3 | 23.5 |
| **MS** |  |  |  |  |  |  |
| *Osteoarthritis* | 23.5 | 40.1 | 50.0 | 45.3 | 44.0 | 37.3 |
| *Osteoporosis* | 23.5 | 31.4 | 41.3 | 54.0 | 66.4 | 90.0 |
| **Sensory** |  |  |  |  |  |  |
| *Hearing* | 7.4 | 9.5 | 13.5 | 27.0 | 53.9 | 54.9 |
| *Eye* | 4.4 | 9.5 | 25.3 | 61.5 | 82.5 | 94.1 |
| **CNS** |  |  |  |  |  |  |
| *Depression* | 25.0 | 16.1 | 8.3 | 9.4 | 14.7 | 13.7 |
| *Parkinson's* | 0 | 0 | 0.87 | 0.4 | 2.1 | 0 |
| *Cognitive Impairment* | 0 | 0 | 0 | 0.4 | 0.7 | 17.7 |
| **Cancer** | 5.9 | 5.1 | 7.8 | 17.6 | 18.2 | 16.7 |
| **Events Over the Course of Study** |  |  |  |  |  |  |
| SBPP < 9.0 | 0 | 1.2 | 1.9 | 5.6 | 20.2 | 66.7 |
| Disability | 0 | 1.5 | 1.3 | 4.7 | 18.3 | 66.7 |
| Geriatric syndrome | 0 | 0 | 1.1 | 34.58 | 61.02 | 86.32 |
| Mortality | 0 | 0.49 | 0.43 | 1.4 | 3.3 | 5.9 |

SD: standard deviation, CVS: cardiovascular system, HTN: hypertension, IHD: ischemic heart disease, CHF: congestive heart failure, PAD: peripheral artery disease, CVA: cerebrovascular accidents, CKD: chronic kidney disease, DM: diabetes mellitus, GID: gastrointestinal disease, COPD: chronic obstructive pulmonary disease, MS: musculoskeletal diseases, CNS: central nervous system, SBPP: a short battery of physical performance.

The prevalence of each organ system morbidity over the 13 years in men and women is depicted in Figure S1.

**Figure S1.**
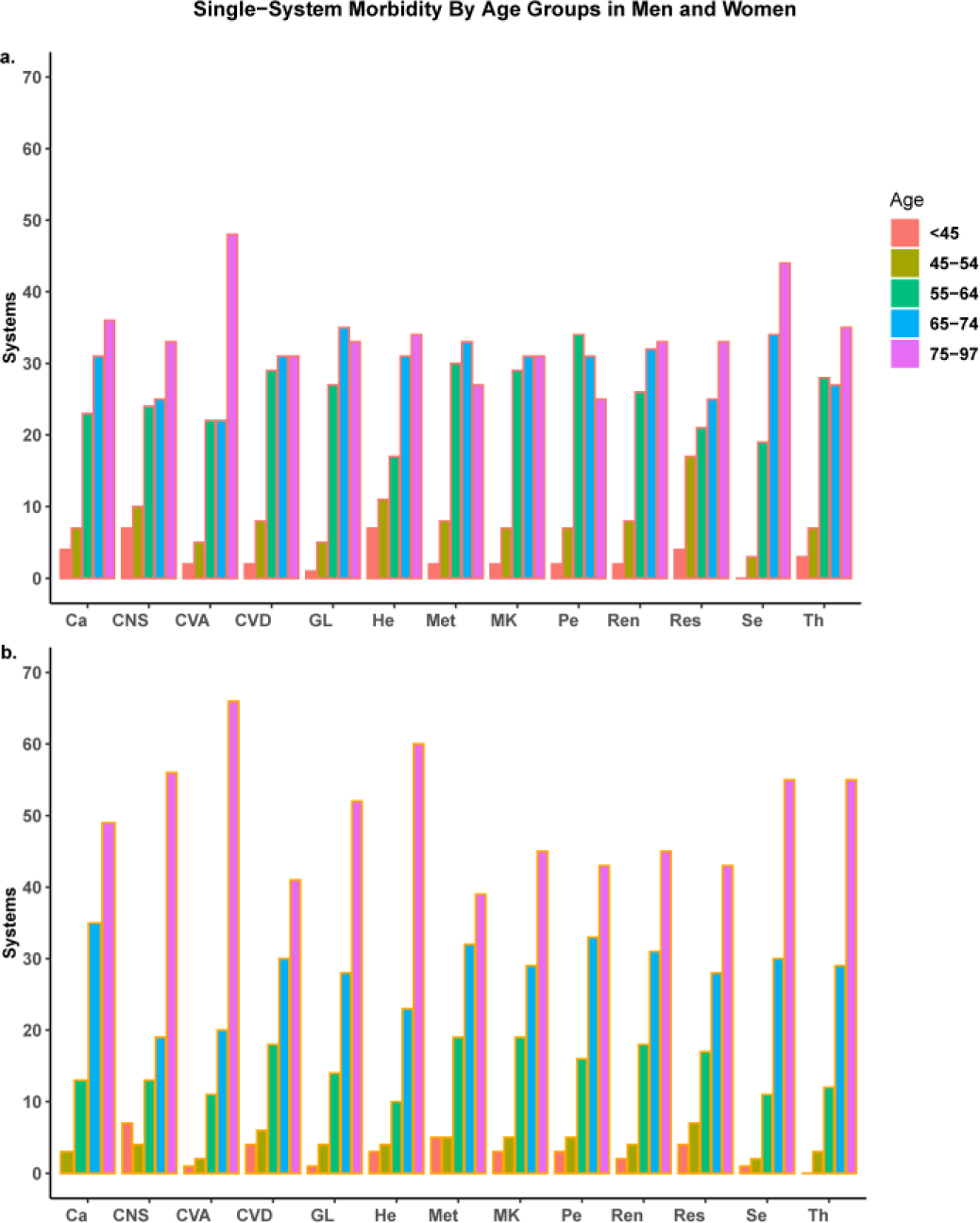
Prevalence of each organ system morbidity by age groups from left to the right: <45, 45–54, 55–64, 65–74, 75–84, >84 years in **a.** men and **b.** women. Renal and cardiovascular systems have the highest prevalence followed by sensory and gastrointestinal, and liver systems. The organ-systems morbidities increased with age. Ca: Cancer, CNS: central nervous system, CVA: cerebrovascular accident, CVS: cardiovascular system, GL: gastrointestinal-liver system, He: hematopoietic system, Met: metabolic system, MK: musculoskeletal system, Pe: periodontitis, Re: renal system, Res: respiratory system, Se: sensory system, Th: dysthyroidism

In the youngest age group (≤ 44 years), >50% of individuals had no morbidity or morbidity in one organ system only. In this age group, participants with cancer frequently have other system morbidities, including CVS, central nervous system (CNS), and renal system (Ren) (Fig. S2a). With increasing age, BODN increases in heterogeneous combinations so that both the number of people with multiple organ-system morbidities and the number of systems with morbidity increase. Particularly, periodontitis, metabolic, kidney, respiratory, and endocrine systems become more frequent after age 55; CVS maintained its increased frequency (Fig. S2c). Renal, sensory (Se), and musculoskeletal (MS) systems appeared more frequently after age 65 (Fig. S2d), while other morbidities such as CVA, cancer, gastrointestinal and liver (GL), and respiratory systems (Res) also further increased. After age 75, most persons showed multiple organ system morbidities, with sensory, CNS, and MS appearing more frequently. CVS, however, was still the most frequent system morbidity in all age groups.

**Figure S2.** Frequency and Combination Patterns for Body Organ Disease Number. The interactive graphs illustrate any existing combined multisystem morbidities in the youngest group (≤ 44 years), >50% of individuals had no morbidity or only one system morbidity. **The dynamic graphs are illustrated in the link below.** https://bodiagesystem.shinyapps.io/BODN/

### Model Assessments

Using posterior predictive checks and LOO-CV model assessment showed high accuracy for both time-1 and time-2 full models, with diagnostics indicating reliable computation (k < 0.5; Table S3, Figs. S3a and S3b; and model convergence Rhat = 1 for all models). Good effective-sample size from chains using MCMC should be >1,000. All models had at least an effective sample size of 2,500. The model assessment showed full models performed better in predicting BODN than their corresponding age-only models (with larger ELPD with diagnostic k<0.5; Table S3).

**Table S3:**
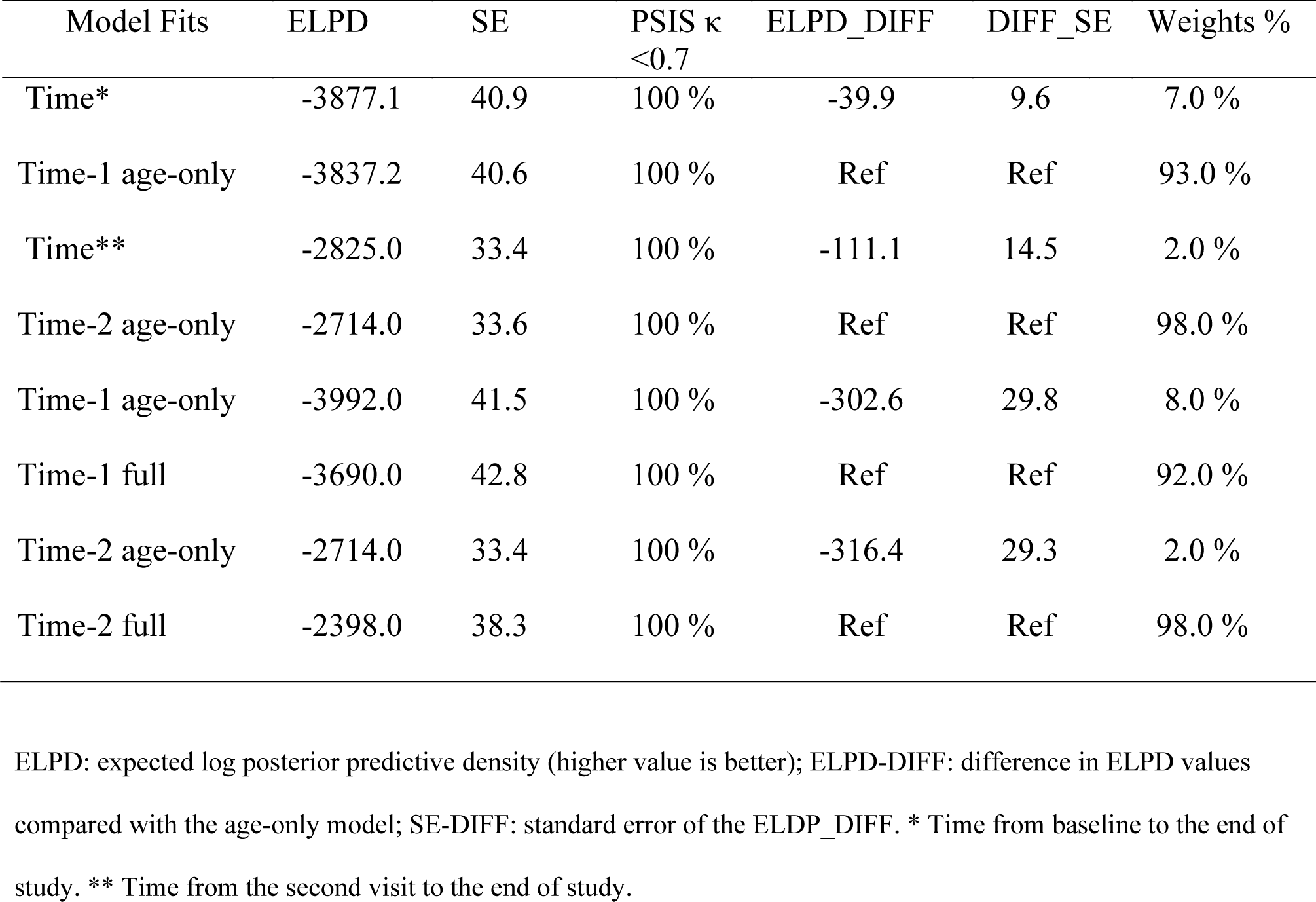
Model Assessments, Model Comparisons, and Model Weights to Predict BODN

**Figure S3.**
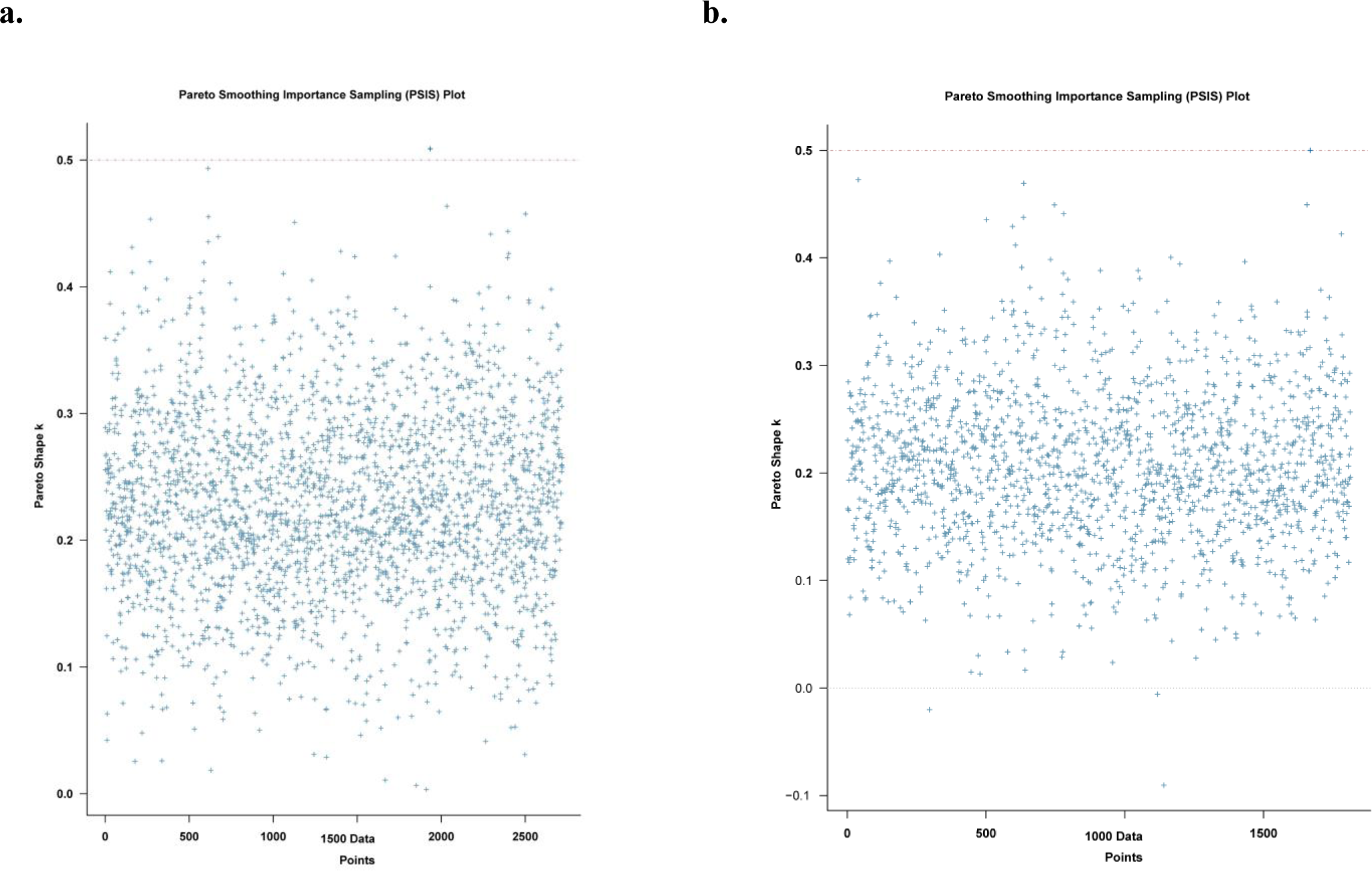
Leave-one-out cross-validation (LOO-CV) for model assessment (with diagnostic k < 0.5). Full models leave-one-out cross-validation in the BLSA at **a.** time-1 and **b.** time-2. Each + represents an individual at each time-point (907×3=2701 observations at time-1, and 907×2=1904 at time-2). Pareto-smoothed importance sampling diagnostic k <0.5 indicates that the LOO-CV computation is reliable and there are no outlier observations.

### In-sample and Out-of-Sample Validation of Body Clock

To validate the BLSA data results, we used the parameters obtained from time-1 BLSA to predict BODN in the InCHIANTI participants as an out-of-sample validation. The external estimates optimally predicted BODN values for InCHIANTI data as illustrated (Fig. S4a). Stacking showed that the full models using InCHIANTI outperformed the age-only models predicting BODN (time-1: 72% versus 28%, time 2: 84% versus 16%). We also used time-1 BLSA parameters (estimates) to create a simulated dataset to compare with the observed data (in-sample validation), showing that the parameters obtained from the model can optimally reproduce the observed BODN (Fig. S4b).

**Figure S4.**
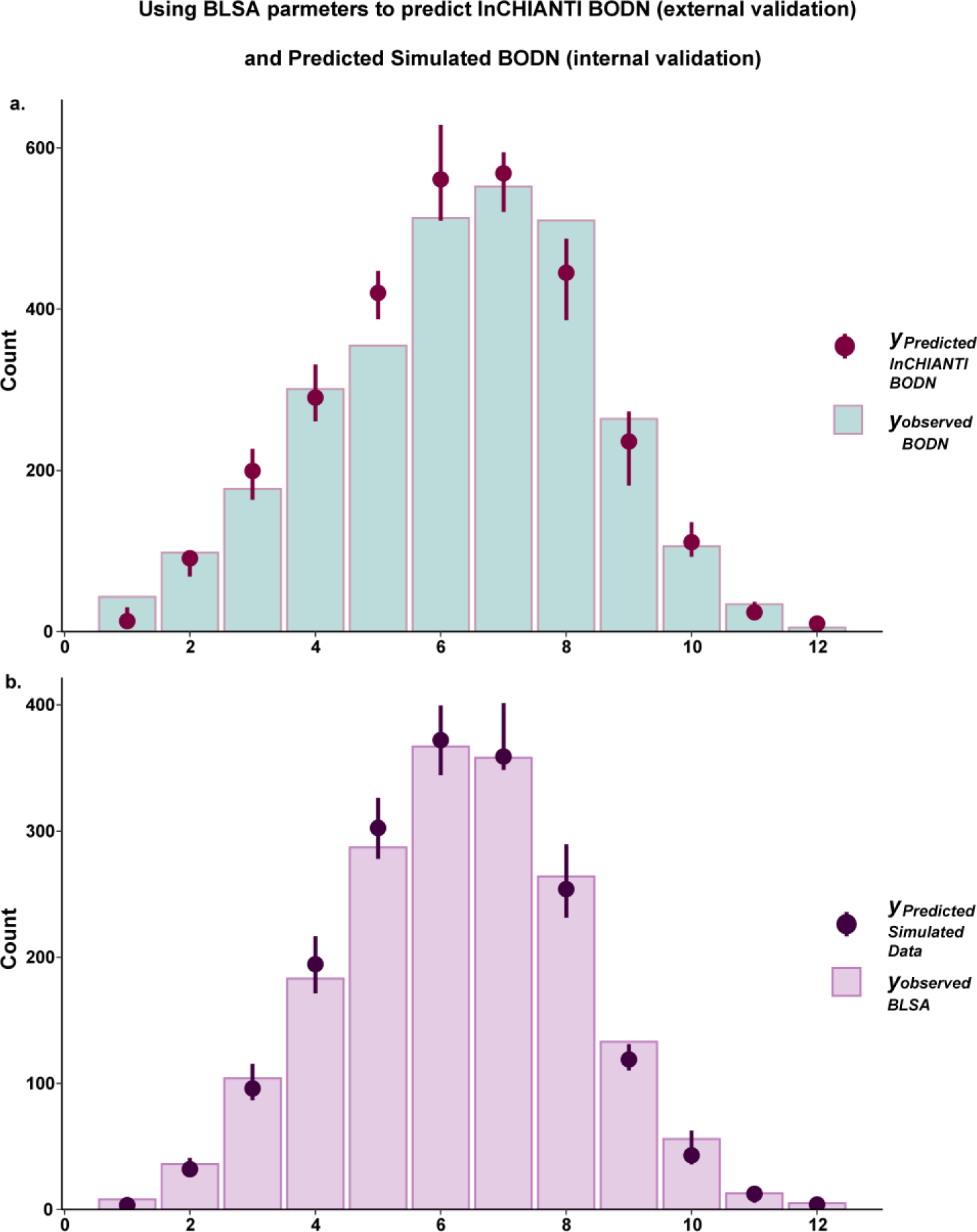
Posterior predictive check (coefficient estimates distribution) of Body Organ Disease Number (BODN). **a.** InCHIANTI using BLSA time-1 lagged parameters (out-of-sample-validation). **b.** Simulated data using BLSA time-1 parameters (In-sample-validation). The predictive check applying distribution of coefficient estimates obtained from BLSA (full model time-1 diseases predicting BODN can be employed to predict BODN in the InCHIANTI dataset (out-of-sample validation). S4b shows that applying the BLSA parameters from the time-1 full model to create a simulated BODN matched the observed BODN in the BLSA dataset (in-sample validation).

**Table S2a.** Time-1 Single Disease Models Predicting Longitudinal BODN Disease

| Disease | Estimate | Estmate.Error | 2.5 % QI | 97.5 % QI |
| --- | --- | --- | --- | --- |
| HTN | 1.48748513 | 0.14244193 | 1.265261 | 1.727344 |
| HTN[L2] | 0.608147942 | 0.02302767 | 0.199595 | 1.180122 |
| HTN[L3] | 0.879337188 | 0.02302767 | 0.400834 | 1.454855 |
| IHD | 1.590701418 | 0.2130871 | 1.236201 | 1.93902 |
| IHD[L2] | 1.405548061 | 0.0164556 | 0.885476 | 1.886387 |
| IHD[L3] | 0.185153357 | 0.0164556 | 0.033556 | 0.550124 |
| CHF | 1.817841746 | 0.19135231 | 1.505468 | 2.132722 |
| CHF[L2] | 0.623780921 | 0.01429419 | 0.335977 | 1.01448 |
| CHF[L3] | 1.194060825 | 0.01429419 | 0.789357 | 1.65676 |
| Arr | 0.745955299 | 0.14010083 | 0.548137 | 1.004203 |
| Arr[L2] | 0.134633477 | 0.01096928 | 0.038362 | 0.329043 |
| Arr[L3] | 0.142959825 | 0.01435785 | 0.030942 | 0.384028 |
| Arr[L4] | 0.160614601 | 0.01634462 | 0.033734 | 0.420957 |
| Arr[L5] | 0.095755181 | 0.01156393 | 0.016202 | 0.303803 |
| Arr[L6] | 0.175775971 | 0.01722097 | 0.036285 | 0.447191 |
| PAD | 3.681348581 | 0.69690847 | 2.527541 | 4.812853 |
| Stroke | 1.658084319 | 0.24100643 | 1.271465 | 2.058649 |
| Stroke[L2] | 0.718711659 | 0.03128027 | 0.294805 | 1.354062 |
| Stroke[L3] | 0.160869135 | 0.01655708 | 0.026049 | 0.537075 |
| Stroke[L4] | 0.753050646 | 0.03122227 | 0.285943 | 1.348193 |
| CKD | 2.543278968 | 0.38526266 | 1.982762 | 3.255165 |
| CKD[L2] | 0.981979037 | 0.02375893 | 0.591656 | 1.616537 |
| CKD[L3] | 0.439258811 | 0.01611823 | 0.216993 | 0.808529 |
| CKD[L4] | 0.386338901 | 0.02596589 | 0.107669 | 0.875411 |
| CKD[L5] | 0.718089258 | 0.04482019 | 0.194285 | 1.476121 |
| DM | 1.100313801 | 0.19663324 | 0.813673 | 1.443534 |
| DM[L2] | 0.198607407 | 0.01352298 | 0.066055 | 0.455984 |
| DM[L3] | 0.062931814 | 0.00808473 | 0.010279 | 0.227045 |
| DM[L4] | 0.295125897 | 0.01868092 | 0.103901 | 0.648871 |
| DM[L5] | 0.117869808 | 0.01239906 | 0.022203 | 0.332538 |
| DM[L6] | 0.400923358 | 0.02374551 | 0.12154 | 0.762908 |
| Lipid | 0.428824981 | 0.21146636 | 0.087281 | 0.786089 |
| Lipid[L2] | 0.195343596 | 0.0478554 | 0.012359 | 0.642082 |
| Lipid[L3] | 0.233481385 | 0.0478554 | 0.015989 | 0.674777 |
| Liver | 1.318365534 | 0.23246367 | 0.986947 | 1.77978 |
| Liver[L2] | 0.542588758 | 0.03254616 | 0.197003 | 1.161116 |
| Liver[L3] | 0.476694279 | 0.03321418 | 0.141961 | 1.07997 |
| Liver[L4] | 0.27348675 | 0.02790752 | 0.05192 | 0.748293 |
| GID | 1.624688808 | 0.18822779 | 1.308383 | 1.939053 |
| GID[L2] | 1.30231338 | 0.02097809 | 0.773181 | 1.836277 |
| GID[L3] | 0.322375427 | 0.02097809 | 0.069348 | 0.793182 |
| COPD | 2.821346678 | 0.45855995 | 2.111571 | 3.614884 |
| COPD[L2] | 1.974075995 | 0.06542288 | 0.989305 | 3.2832 |
| COPD[L3] | 0.847270683 | 0.06542288 | 0.193747 | 1.921252 |
| Asthma | 2.679086915 | 0.5416834 | 1.794785 | 3.587009 |
| Hypoth | 1.150961988 | 0.1483237 | 0.908648 | 1.402722 |
| Hypoth[L2] | 0.708281824 | 0.01762675 | 0.381034 | 1.122354 |
| Hypoth[L3] | 0.207032219 | 0.01618875 | 0.041684 | 0.551865 |
| Hypoth[L4] | 0.206456346 | 0.01601004 | 0.041332 | 0.542585 |
| Hyperth | 2.629559971 | 1.24912886 | 0.601021 | 4.750696 |
| Anemia | 1.499373068 | 0.31865919 | 1.012262 | 2.137788 |
| Anemia[L2] | 1.166531876 | 0.04124908 | 0.551422 | 2.02005 |
| Anemia[L3] | 0.332841192 | 0.04124908 | 0.05575 | 0.973244 |
| Thrombocytopenia | 2.068472599 | 0.65843308 | 0.744881 | 3.324791 |
| OA | 1.369356305 | 0.24714511 | 0.998293 | 1.796632 |
| OA[L2] | 0.91317123 | 0.03097241 | 0.47735 | 1.58459 |
| OA[L3] | 0.456185075 | 0.03097241 | 0.11782 | 0.937543 |
| Osteop | 1.3915404 | 0.18631465 | 1.094002 | 1.7 |
| Osteop[L2] | 0.862914982 | 0.01980504 | 0.498618 | 1.37375 |
| Osteop[L3] | 0.528625419 | 0.01980504 | 0.209952 | 0.925184 |
| Periodontitis | 2.027162054 | 0.30434752 | 1.558269 | 2.530962 |
| Periodontitis[L2] | 1.218530109 | 0.03095982 | 0.702117 | 2.003396 |
| Periodontitis[L3] | 0.808631945 | 0.03095982 | 0.324813 | 1.390574 |
| Hearing | 1.523168862 | 0.18266472 | 1.25482 | 1.849644 |
| Hearing[L2] | 1.016883232 | 0.01693737 | 0.656657 | 1.510456 |
| Hearing[L3] | 0.225454716 | 0.01440523 | 0.05265 | 0.54385 |
| Hearing[L4] | 0.255980065 | 0.01695774 | 0.056576 | 0.624517 |
| Eye | 1.652546546 | 0.20648247 | 1.352296 | 2.044069 |
| Eye[L2] | 0.983275203 | 0.01589571 | 0.647984 | 1.486025 |
| Eye[L3] | 0.084973886 | 0.00696323 | 0.016475 | 0.25369 |
| Eye[L4] | 0.236649432 | 0.01383546 | 0.064188 | 0.521441 |
| Eye[L5] | 0.320243499 | 0.02024355 | 0.074992 | 0.726519 |
| Depression | 1.074648122 | 0.24174024 | 0.6803 | 1.475524 |
| Depression[L2] | 0.6515118 | 0.0480564 | 0.182853 | 1.297759 |
| Depression[L3] | 0.423136322 | 0.0480564 | 0.08196 | 1.078929 |
| Parkinson's | 3.17286003 | 1.36860008 | 1.004209 | 5.669947 |
| Park[L2] | 1.785098423 | 0.28943681 | 0.236803 | 4.948727 |
| Park[L3] | 1.387761607 | 0.28943681 | 0.127736 | 4.332916 |
| CI | 3.281019941 | 0.89599083 | 1.896976 | 4.898348 |
| CI[L2] | 2.311773652 | 0.14338112 | 0.799231 | 4.509921 |
| CI[L3] | 0.969246289 | 0.14338112 | 0.150426 | 2.834585 |
| Cancer | 2.764381135 | 0.41449836 | 2.089097 | 3.439591 |

HTN: Hypertension, IHS: Ischemic heart disease, CHF: Congestive heart failure, Arr: Arrhythmia, PAD: peripheral artery disease, CKD: chronic kidney disease, DM; diabetes mellitus, Lipid; hyperlipidemia, GID: gastrointestinal disease, COPD: Chronic obstructive pulmonary disease, Hypoth: Hypothyroidism, OA: osteoarthritis, osteop: osteoporosis, Park: Parkinson's, CI: cognitive impairment

**Table S2b.**
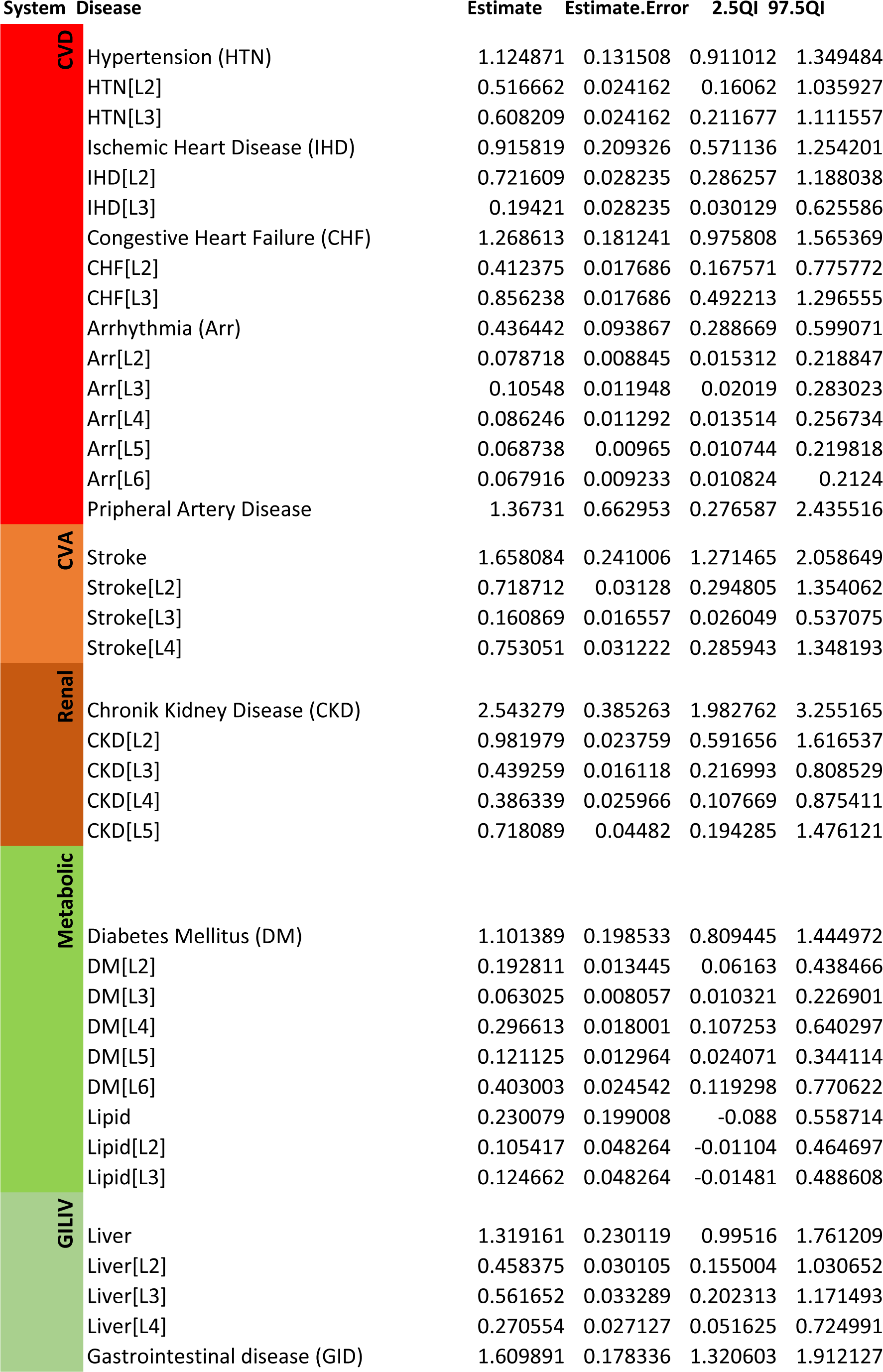
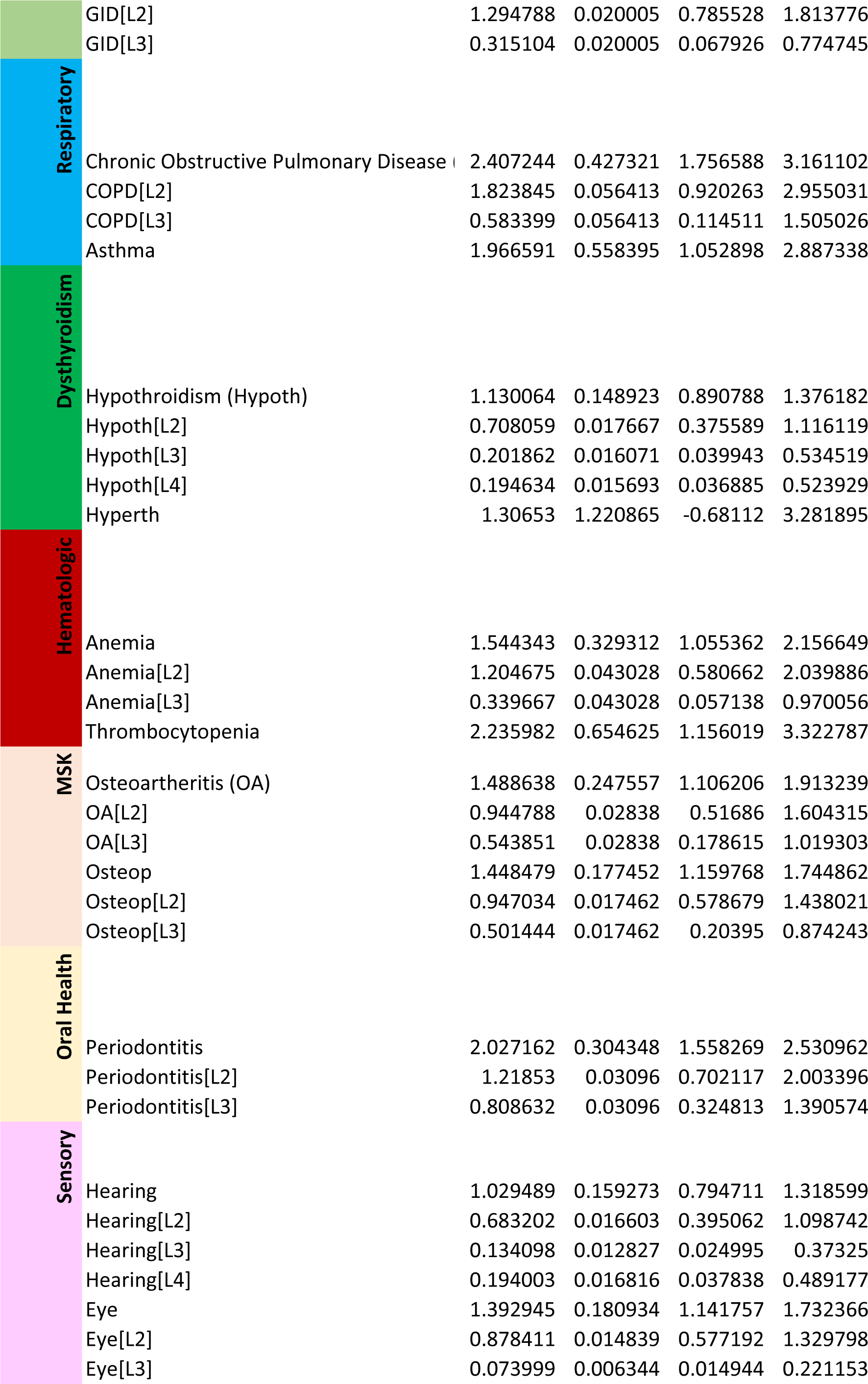
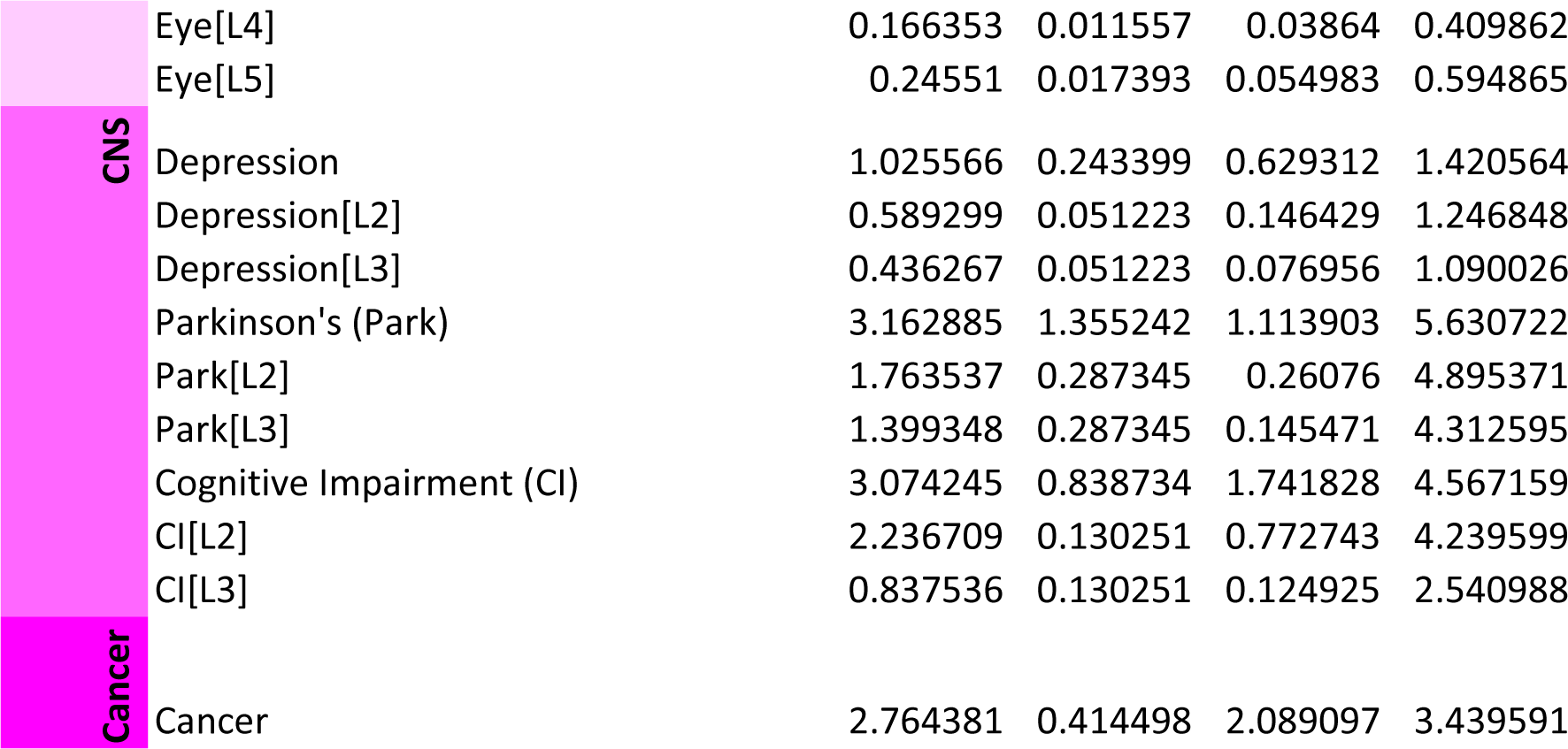
Time-1 Single-System Models Predicting Longitudinal BODN

**Table S2c.** Time-1 Disease Levels of All Organ Systems Predciting Longitudinal BODN

| System | Disease | Estimate | Estimate.Error | 2.5QI | 97.5QI |
| --- | --- | --- | --- | --- | --- |
| CVD | Hypertension (HTN) | 0.4901 | 0.063542 | 0.387963 | 0.5967 |
|  | HTN[L2] | 0.3299 | 0.042782 | 0.261211 | 0.401752 |
|  | HTN[L3] | 0.1601 | 0.02076 | 0.126752 | 0.194948 |
|  | Ischemic Heart Disease (IHD) | 0.2304 | 0.107847 | 0.053816 | 0.408531 |
|  | IHD[L2] | 0.1081 | 0.050608 | 0.025254 | 0.191707 |
|  | IHD[L3] | 0.1223 | 0.057239 | 0.028563 | 0.216825 |
|  | Congestive Heart Failure (CHF) | 0.3216 | 0.074757 | 0.201216 | 0.449982 |
|  | CHF[L2] | 0.2341 | 0.054418 | 0.146471 | 0.327556 |
|  | CHF[L3] | 0.0875 | 0.020339 | 0.054744 | 0.122426 |
|  | Arrhythmia (Arr) | 0.1664 | 0.039274 | 0.102884 | 0.235059 |
|  | Arr[L2] | 0.0697 | 0.016448 | 0.043087 | 0.098441 |
|  | Arr[L3] | 0.027 | 0.006376 | 0.016701 | 0.038158 |
|  | Arr[L4] | 0.0201 | 0.004739 | 0.012413 | 0.028361 |
|  | Arr[L5] | 0.0189 | 0.004461 | 0.011686 | 0.026699 |
|  | Arr[L6] | 0.0206 | 0.004861 | 0.012734 | 0.029094 |
|  | Peripheral Artery Disease | -0.0406 | 0.315954 | -0.5617 | 0.478966 |
| CVA | Stroke | 0.6723 | 0.098464 | 0.512061 | 0.838745 |
|  | Stroke[L2] | 0.3464 | 0.050729 | 0.263817 | 0.432127 |
|  | Stroke[L3] | 0.0935 | 0.013697 | 0.071233 | 0.116678 |
|  | Stroke[L4] | 0.217 | 0.031779 | 0.165268 | 0.270705 |
| Renal | Chronic Kidney Disease (CKD) | 0.9306 | 0.147978 | 0.726341 | 1.222689 |
|  | CKD[L2] | 0.5406 | 0.085965 | 0.421951 | 0.710294 |
|  | CKD[L3] | 0.0708 | 0.011254 | 0.055237 | 0.092984 |
|  | CKD[L4] | 0.0976 | 0.015517 | 0.076166 | 0.128215 |
|  | CKD[L5] | 0.2046 | 0.032536 | 0.1597 | 0.268831 |
| Metabolic | Diabetes Mellitus (DM) | 0.4106 | 0.049724 | 0.337443 | 0.505957 |
|  | DM[L2] | 0.1762 | 0.021342 | 0.144831 | 0.217158 |
|  | DM[L3] | 0.059 | 0.007145 | 0.048489 | 0.072703 |
|  | DM[L4] | 0.0586 | 0.0071 | 0.048183 | 0.072246 |
|  | DM[L5] | 0.0481 | 0.005824 | 0.039522 | 0.059259 |
|  | DM[L6] | 0.0528 | 0.006397 | 0.043411 | 0.06509 |
|  | Lipid | 0.4458 | 0.083246 | 0.314462 | 0.58849 |
|  | Lipid[L2] | 0.3219 | 0.060111 | 0.22707 | 0.424943 |
|  | Lipid[L3] | 0.1239 | 0.023135 | 0.087392 | 0.163547 |
| GILIV | Liver | 0.7769 | 0.133789 | 0.591171 | 1.031132 |
|  | Liver[L2] | 0.4361 | 0.075101 | 0.331848 | 0.578816 |
|  | Liver[L3] | 0.128 | 0.022035 | 0.097366 | 0.169829 |
|  | Liver[L4] | 0.1999 | 0.034431 | 0.15214 | 0.265366 |
|  | Gastrointestinal disease (GI) | 0.9063 | 0.082108 | 0.773708 | 1.047646 |
|  | GID[L2] | 0.7413 | 0.067157 | 0.632825 | 0.856882 |
|  | GID[L3] | 0.165 | 0.014951 | 0.140883 | 0.190764 |
| Respiratory | Chronic Obstructive Pulm | 0.6973 | 0.175453 | 0.414325 | 0.989226 |
|  | COPD[L2] | 0.5629 | 0.141638 | 0.334472 | 0.798574 |
|  | COPD[L3] | 0.1344 | 0.033815 | 0.079852 | 0.190653 |
|  | Asthma | 1.6447 | 0.247988 | 1.241771 | 2.057476 |
| Dysthyroidism | Hypothroidism (Hypoth) | 0.7089 | 0.066144 | 0.604167 | 0.820915 |
|  | Hypoth[L2] | 0.5181 | 0.048342 | 0.441563 | 0.599976 |
|  | Hypoth[L3] | 0.0896 | 0.008359 | 0.076352 | 0.103744 |
|  | Hypoth[L4] | 0.0884 | 0.008245 | 0.075308 | 0.102325 |
|  | Hyperth | -0.0282 | 0.531731 | -0.90793 | 0.838024 |
| Hematologic | Anemia | 0.9656 | 0.18783 | 0.699185 | 1.326615 |
|  | Anemia[L2] | 0.6969 | 0.135558 | 0.504605 | 0.957423 |
|  | Anemia[L3] | 0.2687 | 0.052272 | 0.194581 | 0.369192 |
|  | Thrombocytopenia | 2.1069 | 0.293686 | 1.626414 | 2.604094 |
| MSK | Osteoarthritis (OA) | 0.9236 | 0.114284 | 0.748405 | 1.117593 |
|  | OA[L2] | 0.6574 | 0.081345 | 0.5327 | 0.79548 |
|  | OA[L3] | 0.2662 | 0.032939 | 0.215705 | 0.322113 |
|  | Osteop | 0.5872 | 0.070873 | 0.474086 | 0.707026 |
|  | Osteop[L2] | 0.4998 | 0.060321 | 0.403501 | 0.60176 |
|  | Osteop[L3] | 0.0874 | 0.010552 | 0.070585 | 0.105267 |
| Oral Health | Periodontitis | 1.3705 | 0.11046 | 1.205917 | 1.570483 |
|  | Periodontitis[L2] | 1.1645 | 0.093851 | 1.024595 | 1.334344 |
|  | Periodontitis[L3] | 0.2061 | 0.016609 | 0.181322 | 0.236139 |
| Sensory | Hearing | 0.47 | 0.07944 | 0.348463 | 0.608929 |
|  | Hearing[L2] | 0.2875 | 0.048601 | 0.213188 | 0.37254 |
|  | Hearing[L3] | 0.0939 | 0.015869 | 0.069609 | 0.12164 |
|  | Hearing[L4] | 0.078 | 0.013187 | 0.057844 | 0.101081 |
|  | Eye | 0.6709 | 0.096069 | 0.539163 | 0.854469 |
|  | Eye[L2] | 0.4295 | 0.061503 | 0.345174 | 0.547033 |
|  | Eye[L3] | 0.05 | 0.007157 | 0.040165 | 0.063654 |
|  | Eye[L4] | 0.0386 | 0.00553 | 0.031036 | 0.049186 |
|  | Eye[L5] | 0.1399 | 0.020034 | 0.112436 | 0.178189 |
| CNS | Depression | 0.7574 | 0.104652 | 0.587376 | 0.929958 |
|  | Depression[L2] | 0.4662 | 0.064419 | 0.36156 | 0.572437 |
|  | Depression[L3] | 0.2912 | 0.040233 | 0.225816 | 0.357521 |
|  | Parkinson (Park) | 0.5288 | 0.56304 | -0.48385 | 1.507164 |
|  | Park[L2] | 0.2617 | 0.278659 | -0.23947 | 0.745924 |
|  | Park[L3] | 0.2671 | 0.284381 | -0.24438 | 0.76124 |
|  | Cognitive Impairment (CI) | 1.3213 | 0.353249 | 0.755685 | 1.95154 |
|  | CI[L2] | 1.0165 | 0.271779 | 0.5814 | 1.501454 |
|  | CI[L3] | 0.3047 | 0.08147 | 0.174284 | 0.450086 |
| Cancer | Cancer | 2.3113 | 0.183372 | 2.014284 | 2.61664 |

**Table S2d.** Time-2 Single Disease Models Predicting Longitudinal BODN

| **Disease** | **Estimate** | **Estimate. Error** | **2.5QI** | **97.5 QI** |
| --- | --- | --- | --- | --- |
| HTN | 1.365808 | 0.145896 | 1.084617 | 1.654983 |
| HTN[L2] | 0.457781 | 0.023344 | 0.073561 | 1.118686 |
| HTN[L3] | 0.908027 | 0.023344 | 0.35147 | 1.542738 |
| IHD | 1.622505 | 0.222358 | 1.192195 | 2.065991 |
| IHD[L2] | 1.405864 | 0.017816 | 0.808273 | 2.02548 |
| IHD[L3] | 0.216641 | 0.017816 | 0.023377 | 0.66531 |
| CHF | 1.769312 | 0.180345 | 1.418302 | 2.119335 |
| CHF[L2] | 0.478147 | 0.015519 | 0.153727 | 0.937947 |
| CHF[L3] | 1.291165 | 0.015519 | 0.790608 | 1.889624 |
| Arr | 0.743946 | 0.136673 | 0.506709 | 1.043734 |
| Arr[L2] | 0.139165 | 0.010789 | 0.025686 | 0.375279 |
| Arr[L3] | 0.152824 | 0.014173 | 0.020366 | 0.45125 |
| Arr[L4] | 0.172207 | 0.015866 | 0.021512 | 0.509586 |
| Arr[L5] | 0.118181 | 0.012682 | 0.011569 | 0.390252 |
| Arr[L6] | 0.161569 | 0.015239 | 0.01854 | 0.471411 |
| PAD | 2.978019 | 0.689274 | 1.619879 | 4.338565 |
| Stroke | 1.668384 | 0.229083 | 1.234162 | 2.119765 |
| Stroke[L2] | 0.73642 | 0.028718 | 0.250695 | 1.482205 |
| Stroke[L3] | 0.167642 | 0.015495 | 0.015594 | 0.567612 |
| Stroke[L4] | 0.764322 | 0.029001 | 0.243015 | 1.461221 |
| CKD | 2.442091 | 0.310664 | 1.904067 | 3.104174 |
| CKD[L2] | 1.036799 | 0.01708 | 0.612134 | 1.664554 |
| CKD[L3] | 0.428573 | 0.013215 | 0.188043 | 0.822401 |
| CKD[L4] | 0.464603 | 0.018483 | 0.145306 | 0.955831 |
| CKD[L5] | 0.512116 | 0.028469 | 0.08769 | 1.218533 |
| DM | 0.951331 | 0.12853 | 0.717657 | 1.21484 |
| DM[L2] | 0.234595 | 0.010546 | 0.070654 | 0.508477 |
| DM[L3] | 0.066894 | 0.006142 | 0.006137 | 0.22857 |
| DM[L4] | 0.286248 | 0.012644 | 0.085145 | 0.609971 |
| DM[L5] | 0.148542 | 0.00974 | 0.02146 | 0.391146 |
| DM[L6] | 0.215052 | 0.012681 | 0.037313 | 0.514318 |
| Lipid | 0.558611 | 0.189719 | 0.195398 | 0.942886 |
| Lipid[L2] | 0.304678 | 0.037964 | 0.029306 | 0.851936 |
| Lipid[L3] | 0.253933 | 0.037964 | 0.018848 | 0.801471 |
| Liver | 1.444632 | 0.180459 | 1.134956 | 1.845936 |
| Liver[L2] | 0.621207 | 0.023077 | 0.213619 | 1.269331 |
| Liver[L3] | 0.622685 | 0.023548 | 0.210204 | 1.275429 |
| Liver[L4] | 0.200739 | 0.014479 | 0.022901 | 0.595614 |
| GID | 1.815753 | 0.185757 | 1.455839 | 2.185794 |
| GID[L2] | 1.433338 | 0.019864 | 0.807431 | 2.104678 |
| GID[L3] | 0.382415 | 0.019864 | 0.054027 | 0.973519 |
| COPD | 2.732677 | 0.460804 | 1.880481 | 3.688122 |
| COPD[L2] | 2.000727 | 0.057043 | 0.884057 | 3.48998 |
| COPD[L3] | 0.73195 | 0.057043 | 0.101028 | 1.954252 |
| Asthma | 2.469325 | 0.561051 | 1.364427 | 3.581968 |
| Hypoth | 1.235864 | 0.149337 | 0.9413 | 1.530367 |
| Hypoth[L2] | 0.603571 | 0.018269 | 0.234759 | 1.117869 |
| Hypoth[L3] | 0.317071 | 0.019663 | 0.042818 | 0.821795 |
| Hypoth[L4] | 0.315221 | 0.019071 | 0.043257 | 0.806556 |
| Hyperth | 2.122435 | 1.482759 | -0.77767 | 5.092417 |
| Anemia | 1.591337 | 0.322833 | 1.017547 | 2.288061 |
| Anemia[L2] | 1.185091 | 0.040385 | 0.489339 | 2.181544 |
| Anemia[L3] | 0.406246 | 0.040385 | 0.04737 | 1.187731 |
| Thrombocytopenia | 2.890611 | 0.634849 | 1.659163 | 4.127576 |
| OA | 1.165759 | 0.182332 | 0.81344 | 1.534836 |
| OA[L2] | 0.82754 | 0.020706 | 0.393912 | 1.416313 |
| OA[L3] | 0.338219 | 0.020706 | 0.062815 | 0.791585 |
| Osteop | 1.165759 | 0.182332 | 0.81344 | 1.534836 |
| Osteop[L2] | 0.82754 | 0.020706 | 0.393912 | 1.416313 |
| Osteop[L3] | 0.338219 | 0.020706 | 0.062815 | 0.791585 |
| Periodontitis | 1.871257 | 0.26831 | 1.368994 | 2.407186 |
| Periodontitis[L2] | 1.270717 | 0.028206 | 0.667175 | 2.151126 |
| Periodontitis[L3] | 0.60054 | 0.028206 | 0.145624 | 1.234051 |
| Hearing | 1.559496 | 0.215671 | 1.17085 | 2.014039 |
| Hearing[L2] | 0.938724 | 0.020281 | 0.50103 | 1.595718 |
| Hearing[L3] | 0.286431 | 0.018466 | 0.045657 | 0.732353 |
| Hearing[L4] | 0.334341 | 0.021373 | 0.047961 | 0.841117 |
| Eye | 1.666392 | 0.192839 | 1.328692 | 2.081217 |
| Eye[L2] | 1.035007 | 0.013648 | 0.647295 | 1.591406 |
| Eye[L3] | 0.084664 | 0.006026 | 0.008998 | 0.26293 |
| Eye[L4] | 0.273197 | 0.012249 | 0.060557 | 0.606492 |
| Eye[L5] | 0.273524 | 0.015775 | 0.038655 | 0.704151 |
| Depression | 1.481503 | 0.258922 | 0.97758 | 1.977831 |
| Depression[L2] | 0.783133 | 0.047483 | 0.168334 | 1.715752 |
| Depression[L3] | 0.69837 | 0.047483 | 0.129537 | 1.63726 |
| Parkinson | 2.572778 | 0.771707 | 1.076916 | 4.076622 |
| Park[L2] | 1.501403 | 0.157381 | 0.171917 | 3.774496 |
| Park[L3] | 1.071375 | 0.157381 | 0.079812 | 3.425835 |
| CI | 3.001057 | 0.633862 | 1.869988 | 4.33048 |
| CI[L2] | 2.11621 | 0.08822 | 0.7863 | 4.081947 |
| CI[L3] | 0.884847 | 0.08822 | 0.107322 | 2.509583 |
| Cancer | 2.818 | 0.413636 | 2.034214 | 3.641436 |

HTN: Hypertension, IHS: Ischemic heart disease, CHF: Congestive heart failure, Arr: Arrhythmia, PAD: peripheral artery disease, CKD: chronic kidney disease, DM; diabetes mellitus, Lipid; hyperlipidemia, GID: gastrointestinal disease, COPD: Chronic obstructive pulmonary disease, Hypoth: Hypothyroidism, OA: osteoarthritis, osteop: osteoporosis, Park: Parkinson's, CI: cognitive impairment

**Table S2e.** Time-2 Single-System Models Predicting Longitudinal BODN

| System | Disease | Estimate | Estimate. Error | 2.5QI | 97.5QI |
| --- | --- | --- | --- | --- | --- |
| CVD | Hypertension (HTN) | 1.018357 | 0.143377 | 0.73696 | 1.301482 |
|  | HTN[L2] | 0.418161 | 0.026729 | 0.066469 | 1.035602 |
|  | HTN[L3] | 0.600197 | 0.026729 | 0.150554 | 1.184096 |
|  | Ischemic Heart Disease (IHD) | 1.000125 | 0.216349 | 0.5789 | 1.429937 |
|  | IHD[L2] | 0.729253 | 0.03174 | 0.227183 | 1.362437 |
|  | IHD[L3] | 0.270872 | 0.03174 | 0.027327 | 0.868773 |
|  | Congestive Heart Failure (CHF) | 1.315361 | 0.17528 | 0.9752 | 1.662775 |
|  | CHF[L2] | 0.334378 | 0.017722 | 0.07355 | 0.777206 |
|  | CHF[L3] | 0.980983 | 0.017722 | 0.519377 | 1.537368 |
|  | Arrhythmia (Arr) | 0.467016 | 0.103864 | 0.271467 | 0.683637 |
|  | Arr[L2] | 0.088638 | 0.009675 | 0.010178 | 0.269983 |
|  | Arr[L3] | 0.108507 | 0.012374 | 0.011571 | 0.3402 |
|  | Arr[L4] | 0.101532 | 0.012474 | 0.009 | 0.334287 |
|  | Arr[L5] | 0.090284 | 0.011443 | 0.008163 | 0.305801 |
|  | Arr[L6] | 0.078056 | 0.009979 | 0.006722 | 0.263613 |
|  | Peripheral Artery Disease | 1.10438 | 0.631697 | -0.1545 | 2.346295 |
| CVA | Stroke | 1.668384 | 0.229083 | 1.234162 | 2.119765 |
|  | Stroke[L2] | 0.73642 | 0.028718 | 0.250695 | 1.482205 |
|  | Stroke[L3] | 0.167642 | 0.015495 | 0.015594 | 0.567612 |
|  | Stroke[L4] | 0.764322 | 0.029001 | 0.243015 | 1.461221 |
| Renal | Chronic Kidney Disease (CKD) | 2.442091 | 0.310664 | 1.904067 | 3.104174 |
|  | CKD[L2] | 1.036799 | 0.01708 | 0.612134 | 1.664554 |
|  | CKD[L3] | 0.428573 | 0.013215 | 0.188043 | 0.822401 |
|  | CKD[L4] | 0.464603 | 0.018483 | 0.145306 | 0.955831 |
|  | CKD[L5] | 0.512116 | 0.028469 | 0.08769 | 1.218533 |
| Metabolic | Diabetes Mellitus (DM) | 0.941733 | 0.127327 | 0.707474 | 1.207974 |
|  | DM[L2] | 0.220121 | 0.010625 | 0.057444 | 0.491319 |
|  | DM[L3] | 0.065162 | 0.006047 | 0.005998 | 0.229004 |
|  | DM[L4] | 0.280684 | 0.012822 | 0.080813 | 0.611516 |
|  | DM[L5] | 0.157705 | 0.010227 | 0.023789 | 0.407973 |
|  | DM[L6] | 0.21806 | 0.012769 | 0.035544 | 0.516079 |
|  | Lipid | 0.306806 | 0.183832 | -0.05596 | 0.668182 |
|  | Lipid[L2] | 0.16523 | 0.03903 | -0.00713 | 0.60988 |
|  | Lipid[L3] | 0.141576 | 0.03903 | -0.00488 | 0.583097 |
| GILIV | Liver | 1.38194 | 0.18042 | 1.073583 | 1.783083 |
|  | Liver[L2] | 0.514618 | 0.022064 | 0.159046 | 1.12239 |
|  | Liver[L3] | 0.661507 | 0.023015 | 0.248112 | 1.29947 |
|  | Liver[L4] | 0.205815 | 0.014926 | 0.024015 | 0.595372 |
|  | Gastrointestinal disease (GID) | 1.689544 | 0.174246 | 1.351319 | 2.031869 |
|  | GID[L2] | 1.3655 | 0.017146 | 0.796917 | 1.961903 |
|  | GID[L3] | 0.324044 | 0.017146 | 0.046532 | 0.833609 |
| Respiratory | Chronic Obstructive Pulmonary | 2.42372 | 0.44821 | 1.591579 | 3.332334 |
|  | COPD[L2] | 1.837564 | 0.055401 | 0.780943 | 3.194109 |
|  | COPD[L3] | 0.586156 | 0.055401 | 0.066018 | 1.697252 |
|  | Asthma | 1.658557 | 0.565445 | 0.553314 | 2.754458 |
| Dysthyroidism | Hypothyroidism (Hypoth) | 1.224121 | 0.148128 | 0.93947 | 1.51975 |
|  | Hypoth[L2] | 0.601929 | 0.018744 | 0.229635 | 1.11939 |
|  | Hypoth[L3] | 0.312949 | 0.019576 | 0.039265 | 0.821059 |
|  | Hypoth[L4] | 0.309242 | 0.019058 | 0.04149 | 0.805626 |
|  | Hyperth | 1.728464 | 1.441883 | -1.09406 | 4.554366 |
| Hematologic | Anemia | 1.547565 | 0.321944 | 0.974908 | 2.23388 |
|  | Anemia[L2] | 1.13496 | 0.041509 | 0.455607 | 2.129547 |
|  | Anemia[L3] | 0.412605 | 0.041509 | 0.045533 | 1.189913 |
|  | Thrombocytopenia | 2.670039 | 0.654878 | 1.412888 | 3.992609 |
| MSK | Osteoarthritis (OA) | 1.345266 | 0.219338 | 0.94882 | 1.802361 |
|  | OA[L2] | 1.008486 | 0.023515 | 0.506721 | 1.699702 |
|  | OA[L3] | 0.33678 | 0.023515 | 0.054043 | 0.839804 |
|  | Osteop | 1.189374 | 0.176104 | 0.850037 | 1.545254 |
|  | Osteop[L2] | 0.896277 | 0.01855 | 0.454305 | 1.453848 |
|  | Osteop[L3] | 0.293096 | 0.01855 | 0.050282 | 0.719389 |
| Oral Health | Periodontitis | 1.871257 | 0.26831 | 1.368994 | 2.407186 |
|  | Periodontitis[L2] | 1.270717 | 0.028206 | 0.667175 | 2.151126 |
|  | Periodontitis[L3] | 0.60054 | 0.028206 | 0.145624 | 1.234051 |
| Sensory | Hearing | 1.139536 | 0.193769 | 0.789897 | 1.544018 |
|  | Hearing[L2] | 0.639806 | 0.021346 | 0.282885 | 1.209254 |
|  | Hearing[L3] | 0.189005 | 0.01728 | 0.021745 | 0.565522 |
|  | Hearing[L4] | 0.310724 | 0.023063 | 0.044807 | 0.784402 |
|  | Eye | 1.469936 | 0.1774 | 1.159481 | 1.856872 |
|  | Eye[L2] | 0.939982 | 0.013364 | 0.576022 | 1.467763 |
|  | Eye[L3] | 0.076834 | 0.005816 | 0.008069 | 0.247109 |
|  | Eye[L4] | 0.20955 | 0.01113 | 0.037963 | 0.504977 |
|  | Eye[L5] | 0.24357 | 0.014698 | 0.033434 | 0.63251 |
| CNS | Depression | 1.348211 | 0.257022 | 0.856412 | 1.861212 |
|  | Depression[L2] | 0.692507 | 0.048249 | 0.132127 | 1.624393 |
|  | Depression[L3] | 0.655704 | 0.048249 | 0.108969 | 1.574065 |
|  | Parkinson (Park) | 2.070603 | 0.761152 | 0.556259 | 3.581636 |
|  | Park[L2] | 1.185202 | 0.159305 | 0.083112 | 3.306083 |
|  | Park[L3] | 0.885401 | 0.159305 | 0.042796 | 3.046493 |
|  | Cognitive Impairment (CI) | 2.616761 | 0.613695 | 1.484476 | 3.909597 |
|  | CI[L2] | 1.792295 | 0.089525 | 0.574103 | 3.670461 |
|  | CI[L3] | 0.824466 | 0.089525 | 0.0908 | 2.397608 |
| Cancer | Cancer | 2.818 | 0.413636 | 2.034214 | 3.641436 |

**Table S2f.** Time-2 Disease Levels of All Organ Systems Predicting Longitudinal BODN

| System | Disease | Estimate | Estimate.Error | 2.5QI | 97.5QI |
| --- | --- | --- | --- | --- | --- |
| CVD | Hypertension (HTN) | 0.379916 | 0.059904 | 0.262583 | 0.497771 |
|  | HTN[L2] | 0.232224 | 0.011382 | 0.056743 | 0.462956 |
|  | HTN[L3] | 0.147692 | 0.011382 | 0.018366 | 0.390204 |
|  | Ischemic Heart Disease (IHD) | 0.182975 | 0.099149 | -0.01024 | 0.375307 |
|  | IHD[L2] | 0.101525 | 0.021356 | -0.00128 | 0.346461 |
|  | IHD[L3] | 0.081449 | 0.021356 | -0.00079 | 0.328557 |
|  | Congestive Heart Failure (CHF) | 0.269525 | 0.073366 | 0.126922 | 0.416864 |
|  | CHF[L2] | 0.151511 | 0.013051 | 0.02594 | 0.372765 |
|  | CHF[L3] | 0.118014 | 0.013051 | 0.013427 | 0.331668 |
|  | Arrhythmia (Arr) | 0.206914 | 0.036899 | 0.137996 | 0.283298 |
|  | Arr[L2] | 0.107797 | 0.003952 | 0.042722 | 0.206691 |
|  | Arr[L3] | 0.030836 | 0.003135 | 0.003271 | 0.098 |
|  | Arr[L4] | 0.021089 | 0.002399 | 0.001962 | 0.073197 |
|  | Arr[L5] | 0.02183 | 0.002435 | 0.001972 | 0.074123 |
|  | Arr[L6] | 0.025362 | 0.002743 | 0.002472 | 0.084984 |
|  | Peripheral Artery Disease | -0.04873 | 0.275327 | -0.58769 | 0.491798 |
| CVA | Stroke | 0.781978 | 0.091318 | 0.61067 | 0.966722 |
|  | Stroke[L2] | 0.4037 | 0.011461 | 0.163408 | 0.73768 |
|  | Stroke[L3] | 0.142072 | 0.009589 | 0.017279 | 0.410499 |
|  | Stroke[L4] | 0.236206 | 0.010205 | 0.053314 | 0.50154 |
| Renal | Chronic Kidney Disease (CKD) | 0.842725 | 0.139055 | 0.605728 | 1.143257 |
|  | CKD[L2] | 0.518513 | 0.01307 | 0.270452 | 0.921447 |
|  | CKD[L3] | 0.043151 | 0.003882 | 0.005324 | 0.131483 |
|  | CKD[L4] | 0.056048 | 0.005493 | 0.006196 | 0.181499 |
|  | CKD[L5] | 0.225013 | 0.015071 | 0.035734 | 0.536295 |
| Metabolic | Diabetes Mellitus (DM) | 0.386649 | 0.038848 | 0.315486 | 0.467351 |
|  | DM[L2] | 0.217345 | 0.003384 | 0.123763 | 0.342049 |
|  | DM[L3] | 0.049183 | 0.00275 | 0.00679 | 0.135443 |
|  | DM[L4] | 0.035684 | 0.002165 | 0.004144 | 0.104461 |
|  | DM[L5] | 0.052721 | 0.002552 | 0.008262 | 0.127662 |
|  | DM[L6] | 0.031715 | 0.001949 | 0.003493 | 0.093711 |
|  | Lipid | 0.498728 | 0.068279 | 0.365821 | 0.636966 |
|  | Lipid[L2] | 0.440733 | 0.004888 | 0.261312 | 0.626467 |
|  | Lipid[L3] | 0.057995 | 0.004888 | 0.006029 | 0.18197 |
| GILIV | Liver | 0.807865 | 0.102893 | 0.632982 | 1.035853 |
|  | Liver[L2] | 0.498323 | 0.010576 | 0.266522 | 0.848682 |
|  | Liver[L3] | 0.154523 | 0.008837 | 0.028778 | 0.387139 |
|  | Liver[L4] | 0.155019 | 0.009234 | 0.024971 | 0.391196 |
|  | Gastrointestinal disease (GID) | 0.971765 | 0.078902 | 0.821113 | 1.127105 |
|  | GID[L2] | 0.797555 | 0.00683 | 0.521177 | 1.087994 |
|  | GID[L3] | 0.17421 | 0.00683 | 0.028493 | 0.411709 |
| Respiratory | Chronic Obstructive Pulmonary Disease (COPD) | 1.054557 | 0.180949 | 0.720707 | 1.43213 |
|  | COPD[L2] | 0.854204 | 0.018734 | 0.418053 | 1.387046 |
|  | COPD[L3] | 0.200353 | 0.018734 | 0.022688 | 0.60141 |
|  | Asthma | 1.481823 | 0.236173 | 1.018359 | 1.940204 |
| Dysthyroidism | Hypothyroidism (Hypoth) | 0.809764 | 0.060403 | 0.692728 | 0.929966 |
|  | Hypoth[L2] | 0.591887 | 0.004773 | 0.394687 | 0.817915 |
|  | Hypoth[L3] | 0.127868 | 0.004774 | 0.019065 | 0.309397 |
|  | Hypoth[L4] | 0.090009 | 0.003882 | 0.011854 | 0.242219 |
|  | Hyperth | 1.479465 | 0.571852 | 0.35318 | 2.604114 |
| Hematologic | Anemia | 0.872939 | 0.114071 | 0.665332 | 1.11532 |
|  | Anemia[L2] | 0.748036 | 0.009246 | 0.449185 | 1.092055 |
|  | Anemia[L3] | 0.124903 | 0.009246 | 0.013878 | 0.362335 |
|  | Thrombocytopenia | 1.77794 | 0.275766 | 1.237905 | 2.330068 |
| MSK | Osteoarthritis (OA) | 0.732732 | 0.092765 | 0.565253 | 0.929261 |
|  | OA[L2] | 0.586542 | 0.008107 | 0.353203 | 0.887675 |
|  | OA[L3] | 0.146191 | 0.008107 | 0.025296 | 0.348605 |
|  | Osteop | 0.632623 | 0.065177 | 0.507485 | 0.764525 |
|  | Osteop[L2] | 0.56705 | 0.003875 | 0.385834 | 0.75313 |
|  | Osteop[L3] | 0.065573 | 0.003875 | 0.007563 | 0.183267 |
| Oral Health | Periodontitis | 1.326301 | 0.095104 | 1.152712 | 1.523607 |
|  | Periodontitis[L2] | 1.18852 | 0.004988 | 0.901671 | 1.495978 |
|  | Periodontitis[L3] | 0.137781 | 0.004988 | 0.020904 | 0.331816 |
| Sensory | Hearing | 0.461604 | 0.090482 | 0.297582 | 0.651263 |
|  | Hearing[L2] | 0.233712 | 0.01072 | 0.087032 | 0.488882 |
|  | Hearing[L3] | 0.085783 | 0.009331 | 0.008942 | 0.274269 |
|  | Hearing[L4] | 0.14211 | 0.011744 | 0.020134 | 0.362675 |
|  | Eye | 0.700673 | 0.081251 | 0.563311 | 0.878643 |
|  | Eye[L2] | 0.501037 | 0.006122 | 0.318461 | 0.753921 |
|  | Eye[L3] | 0.039964 | 0.002679 | 0.004938 | 0.1185 |
|  | Eye[L4] | 0.04458 | 0.00306 | 0.005305 | 0.133565 |
|  | Eye[L5] | 0.115092 | 0.006577 | 0.016767 | 0.293015 |
| CNS | Depression | 1.049915 | 0.108762 | 0.840383 | 1.265493 |
|  | Depression[L2] | 0.465469 | 0.014725 | 0.158678 | 0.911871 |
|  | Depression[L3] | 0.584447 | 0.014725 | 0.234831 | 1.026547 |
|  | Parkinson (Park) | 0.764393 | 0.306402 | 0.164649 | 1.371399 |
|  | Park[L2] | 0.394865 | 0.066681 | 0.017924 | 1.241619 |
|  | Park[L3] | 0.369527 | 0.066681 | 0.015581 | 1.222107 |
|  | Cognitive Impairment (CI) | 1.238126 | 0.254549 | 0.774515 | 1.770587 |
|  | CI[L2] | 0.874406 | 0.034934 | 0.329794 | 1.676514 |
|  | CI[L3] | 0.36372 | 0.034934 | 0.041151 | 1.016659 |
| Cancer | Cancer | 2.418129 | 0.178807 | 2.07605 | 2.774339 |

## Notes

### Competing Interest Statement

The authors have declared no competing interest.

### Funding Statement

Shabnam Salimi is funded by NIA-B K01AG059898
Shabnam Salimi is a special volunteer at NIA/IRP.

### Author Declarations

All study participants of BLAS and InCHIANTI studies have signed the informed consent form to use the collected data for the research after their recruitment. Both BLSA and InCHIANTI studies have an overall IRB approval at NIA/IRP to use the data and for the present work. The ethics oversight the collection of the original data at the time of data collection. The current work is the secondary data analyses and IRB regards the present study in its entirety.

### Summary of Updates

The coauthors' first name and last name order needed to be corrected. Now the name is First name and last name in order.The second author's first name is Aki and the last name is Vehtari.

## References

1. Rocca, W.A., et al. Prevalence of multimorbidity in a geographically defined American population: patterns by age, sex, and race/ethnicity. Mayo Clin Proc 89, 1336–1349 (2014).

2. Salive, M.E. Multimorbidity in older adults. Epidemiol Rev 35, 75–83 (2013).

3. Cesari, M., Perez-Zepeda, M.U. & Marzetti, E. Frailty and Multimorbidity: Different Ways of Thinking About Geriatrics. J Am Med Dir Assoc 18, 361–364 (2017).

4. St Sauver, J.L., et al. Risk of developing multimorbidity across all ages in an historical cohort study: differences by sex and ethnicity. BMJ Open 5, e006413 (2015).

5. Fabbri, E., et al. Aging and Multimorbidity: New Tasks, Priorities, and Frontiers for Integrated Gerontological and Clinical Research. J Am Med Dir Assoc 16, 640–647 (2015).

6. Ferrucci, L., et al. Measuring biological aging in humans: A quest. Aging Cell 19, e13080 (2020).

7. Kennedy, B.K., et al. Geroscience: linking aging to chronic disease. Cell 159, 709–713 (2014).

8. Fabbri, E., et al. Aging and the burden of multimorbidity: associations with inflammatory and anabolic hormonal biomarkers. J Gerontol A Biol Sci Med Sci 70, 63–70 (2015).

9. Wei, M.Y., Kabeto, M.U., Langa, K.M. & Mukamal, K.J. Multimorbidity and Physical and Cognitive Function: Performance of a New Multimorbidity-Weighted Index. J Gerontol A Biol Sci Med Sci 73, 225–232 (2018).

10. van Walraven, C., Austin, P.C., Jennings, A., Quan, H. & Forster, A.J. A modification of the Elixhauser comorbidity measures into a point system for hospital death using administrative data. Med Care 47, 626–633 (2009).

11. Moore, B.J., White, S., Washington, R., Coenen, N. & Elixhauser, A. Identifying Increased Risk of Readmission and In-hospital Mortality Using Hospital Administrative Data: The AHRQ Elixhauser Comorbidity Index. Med Care 55, 698–705 (2017).

12. Liu, Z., et al. A new aging measure captures morbidity and mortality risk across diverse subpopulations from NHANES IV: A cohort study. PLoS Med 15, e1002718 (2018).

13. Belsky, D.W., et al. Quantification of biological aging in young adults. Proc Natl Acad Sci U S A 112, E4104–4110 (2015).

14. Rockwood, K., McMillan, M., Mitnitski, A. & Howlett, S.E. A Frailty Index Based on Common Laboratory Tests in Comparison With a Clinical Frailty Index for Older Adults in Long-Term Care Facilities. J Am Med Dir Assoc 16, 842–847 (2015).

15. Li, X., et al. Longitudinal trajectories, correlations and mortality associations of nine biological ages across 20-years follow-up. Elife 9(2020).

16. Rockwood, K., Andrew, M. & Mitnitski, A. A comparison of two approaches to measuring frailty in elderly people. J Gerontol A Biol Sci Med Sci 62, 738–743 (2007).

17. Rutenberg, A.D., Mitnitski, A.B., Farrell, S.G. & Rockwood, K. Unifying aging and frailty through complex dynamical networks. Exp Gerontol 107, 126–129 (2018).

18. Pilotto, A., Addante, F., D’Onofrio, G., Sancarlo, D. & Ferrucci, L. The Comprehensive Geriatric Assessment and the multidimensional approach. A new look at the older patient with gastroenterological disorders. Best Pract Res Clin Gastroenterol 23, 829–837 (2009).

19. Whitty, C.J.M. & Watt, F.M. Map clusters of diseases to tackle multimorbidity. Nature 579, 494–496 (2020).

20. Vehtari A., Gelman, A., Gabry, J. Practical bayesian model evaluation using leave-one-out cross-validation and WAIC. Statistics and Computing 27, 1413–1432 (2017).

21. Yao, Y., Vehtari, A., Simpson, D. & Gelman, A. Using stacking to average Bayesian predictive distributions. Bayesian Analysis 13, 917–1003 (2018).

22. Brilleman, S. L., Elic, E. M., Novik, J. B. & Wolfe, R. Bayesian Survival Analysis Using the rstanarm R Package. arXiv:2002.09633 (2020).

23. Guralnik, J.M., et al. A short physical performance battery assessing lower extremity function: association with self-reported disability and prediction of mortality and nursing home admission. J Gerontol 49, M85–94 (1994).

24. Guralnik, J.M., LaCroix, A.Z., Branch, L.G., Kasl, S.V. & Wallace, R.B. Morbidity and disability in older persons in the years prior to death. Am J Public Health 81, 443–447 (1991).

25. Inouye, S.K., Studenski, S., Tinetti, M.E. & Kuchel, G.A. Geriatric syndromes: clinical, research, and policy implications of a core geriatric concept. J Am Geriatr Soc 55, 780–791 (2007).

26. Charlson, M.E., et al. The Charlson comorbidity index is adapted to predict costs of chronic disease in primary care patients. J Clin Epidemiol 61, 1234–1240 (2008).

27. Salvi, F., et al. A manual of guidelines to score the modified cumulative illness rating scale and its validation in acute hospitalized elderly patients. J Am Geriatr Soc 56, 1926–1931 (2008).

28. Fried, L.P., et al. Nonlinear multisystem physiological dysregulation associated with frailty in older women: implications for etiology and treatment. J Gerontol A Biol Sci Med Sci 64, 1049–1057 (2009).

29. Kuo, P.L., et al. A roadmap to build a phenotypic metric of ageing: insights from the Baltimore Longitudinal Study of Aging. J Intern Med 287, 373–394 (2020).

30. Wei, M.Y., Ratz, D. & Mukamal, K.J. Multimorbidity in Medicare Beneficiaries: Performance of an ICD-Coded Multimorbidity-Weighted Index. J Am Geriatr Soc 68, 999–1006 (2020).

31. Marsh, A.P., et al. Hospitalizations During a Physical Activity Intervention in Older Adults at Risk of Mobility Disability: Analyses from the Lifestyle Interventions and Independence for Elders Randomized Clinical Trial. J Am Geriatr Soc 64, 933–943 (2016).

32. Calderon-Larranaga, A., et al. Multimorbidity and functional impairment-bidirectional interplay, synergistic effects and common pathways. J Intern Med 285, 255–271 (2019).

33. Levine, M.E. Modeling the rate of senescence: can estimated biological age predict mortality more accurately than chronological age? J Gerontol A Biol Sci Med Sci 68, 667–674 (2013).

34. Pfeffer, M.A., Shah, A.M. & Borlaug, B.A. Heart Failure With Preserved Ejection Fraction In Perspective. Circ Res 124, 1598–1617 (2019).

35. Klausner, S.C. & Schwartz, A.B. The aging heart. Clin Geriatr Med 1, 119–141 (1985).

36. Monfredi, O. & Boyett, M.R. Sick sinus syndrome and atrial fibrillation in older persons - A view from the sinoatrial nodal myocyte. J Mol Cell Cardiol 83, 88–100 (2015).

37. Menezes, S.T., et al. Hypertension, Prehypertension, and Hypertension Control: Association With Decline in Cognitive Performance in the ELSA-Brasil Cohort. Hypertension 77, 672–681 (2020).

38. Weiner, D.E., et al. Cognitive Function and Kidney Disease: Baseline Data From the Systolic Blood Pressure Intervention Trial (SPRINT). Am J Kidney Dis 70, 357–367 (2017).

39. Rapp, S.R., et al. Effects of intensive versus standard blood pressure control on domain-specific cognitive function: a substudy of the SPRINT randomised controlled trial. Lancet Neurol 19, 899–907 (2020).

40. Yang, M. & Williamson, J. Blood Pressure and Statin Effects on Cognition: a Review. Curr Hypertens Rep 21, 70 (2019).

41. Zampino, M., AlGhatrif, M., Kuo, P.L., Simonsick, E.M. & Ferrucci, L. Longitudinal Changes in Resting Metabolic Rates with Aging Are Accelerated by Diseases. Nutrients 12(2020).

42. Ferrucci, L., et al. Subsystems contributing to the decline in ability to walk: bridging the gap between epidemiology and geriatric practice in the InCHIANTI study. J Am Geriatr Soc 48, 1618–1625 (2000).

43. Bürkner, P. C., Charpentier, E. Ordinal Regression Models in Psychology: A Tutorial. Advances in Methods and Practices in Psychological Science 2, 77–101 (2019).

44. Gabry, J., Simpson, D.,Vehtari, A.,Betancourt, M., Gelman, A. Visualization in Bayesian workflow. Journal of the Royal Statistical Society Series A 182, 389–402 (2019).

45. Gelman, A., Carlin, J. B., Stern, H., Dunson, D. B, Vehtari, A. & Rubin, B. Bayesian Data Analysis, (Chapman & Hall, 2014).

46. McElreath, R. Statistical Rethinking: A Bayesian Course with Examples in R and Stan, (Chapman and Hall/CRC, USA, 2016).

47. Bürkner P. C. Advanced Bayesian Multilevel Modeling with the R Package brms The R Journal, 10(*1*), 395–411 10(2018).

48. Carpenter B, Gelman, A., Hoffman, M.D., Lee, D., Goodrich, B., Betancourt & M., Riddell, A. Stan: A probabilistic programming language. Journal of Statistical Software 76(2017).

49. Stan Development Team. Stan Modeling Language Users Guide.https://mc-stan.org/users/documentation/. (2020).

50. Bürkner, P. C., Charpentier, E. Modelling monotonic effects of ordinal predictors in Bayesian regression models. British Journal of mathematical and Statistical Pscycology (2020).

51. Vehtari, A., Gelman, A., Simpson, D., Carpenter, B. & Bürkner, P.C. Rank-normalization, folding, and localization:An improved R-hat for assessing convergence of MCMC. Bayesian analysis, 1–28 (2020).

52. Vehtari, A., Gelman, A., Gabry, J. Practical bayesian model evaluation using leave-one-out cross-validation and WAIC. Statistics and Computing 27 1413–1432 (2017).

53. Vehtari, A., Simpson, D., Gelman, A., Yao, Y. & Gabry, J. Pareto smoothed importance sampling. arXiv preprint arXiv:1507.02646 (2019).

54. Cichoz-Lach, H., et al. The BARD score and the NAFLD fibrosis score in the assessment of advanced liver fibrosis in nonalcoholic fatty liver disease. Med Sci Monit 18, CR735-740 (2012).

55. Biondi, B., Cappola, A.R. & Cooper, D.S. Subclinical Hypothyroidism: A Review. JAMA 322, 153–160 (2019).

